# Deep latent variable modelling reveals clinically significant subgroups among transfusion recipients

**DOI:** 10.1101/2025.10.29.25338961

**Authors:** Elissa Peltola, Esa Turkulainen, Markus Heinonen, Mikko Arvas, Minna Ilmakunnas, Miika Koskinen

## Abstract

**Background:** Transfusion recipients are a heterogeneous group of patients, yet the identification of these groups has traditionally relied on human-driven univariate analyses and domain knowledge instead of analyzing multivariate characteristics of individuals. Electronic health records (EHR) combined with unsupervised machine learning enables robust, data-driven way for phenotyping patient populations, providing finer-grained view on subgroup characteristics.

**Materials and Methods:** We introduce an extension to the Variational Autoencoder (VAE) framework and apply the model to EHR data of 19,629 adult transfusion recipients. The latent representation of VAEs approximates a low-dimensional manifold of input data, in which patients with similar characteristics are embedded close to one another. The model integrates clustering via a Gaussian Mixture Model (GMM) prior to identify clinically relevant patient subgroups from diagnosis codes, laboratory values and demographics, while simultaneously classifying the type of transfused products. The final clusters are derived with a modified consensus clustering approach.

**Results:** We identified six patient groups with distinct diagnosis, laboratory, demographic, and transfusion profiles. These novel clusters provide a refined characterization of transfusion-related phenotypes, revealing more detailed distinctions among patient subgroups. Our model achieved moderate classification accuracy, with AUROC of 0.879, 0.806 and 0.861, and PR-AUC of 0.448, 0.357 and 0.492 for red blood cells (RBC), plasma and platelets, respectively. Clustering accuracy remains consistent across training and test sets.

**Conclusions:** Data-driven phenotyping of transfusion recipients revealed previously unexplored patient phenotypes differing in multiple characteristics. The model helps to understand the heterogeneous nature of patients requiring transfusion and provides insights on how different blood product profiles shift cluster assignments. These findings underscore the utility of latent variable modelling for population characterization and suggest potential applications in optimizing transfusion strategies as well as blood supply chain management. Validation in external cohorts remains unestablished.

## 1 Introduction

Blood transfusions are commonly administered to bleeding or anemic patients, and those undergoing major surgery [1, 2]. However, identification and characterization of these groups has largely relied on population-level demographics [3–7], frequencies of diagnosis categories or hospital units [3–6, 8–10], pre-transfusion hemoglobin [3, 9], product distributions [4, 8–10], or domain knowledge. While informative, these statistics overlook the heterogeneous nature of patient phenotypes.

Electronic health records (EHR) provide a rich foundation for data-driven phenotyping of patient populations [11]. Instead of relying on domain knowledge and univariate analyses to characterize transfusion recipients, unsupervised machine learning with large-scale data provide means to identify meaningful subgroups through comprehensive representations of patient profiles. Although these methods have been applied to patients receiving certain blood products [12] or suffering from particular diseases [13], the heterogeneity and phenotype characteristics of the broader transfusion recipient population remain poorly understood.

In this study, we introduce an unsupervised machine learning model to identify and characterize patient subgroups and investigate how distributions of transfused products vary between them using high-resolution EHR data from 19,629 adult transfusion recipients. We extend the Variational Autoencoder (VAE) [14] framework with a Gaussian Mixture Model (GMM) prior and a discriminative classification module that incorporates blood product information into the patient subgrouping pipeline. Phenotypes are derived using a modified consensus clustering [15] approach to ensure robustness, while the overall method enables explainability regarding the influence of product information on phenotype formation and detailed characterization of multivariate phenotypic profiles.

## 2 Materials and Methods

### 2.1 Data

Data comprises patient age, sex, body mass index (BMI), ICD-10 codes, laboratory results and transfusion records of adult (≥ 18 years) transfusion recipients treated at HUS Helsinki University Hospital during 2021-2022.

We collected unique ICD-10 codes, expressed as the first three characters, over 4 years preceding the patient’s transfusion. We excluded diagnosis codes for congenital malformations, deformations and chromosomal abnormalities (Q00-Q99), symptoms, signs and abnormal clinical and laboratory findings, not elsewhere classified (R00-R99), and factors influencing health status and contact with health services Z00-Z99 (Z00-Z99). As an exception, Z94 (transplanted organ and tissue status) and Z95 (presence of cardiac and vascular implants and grafts) were included.

For laboratory findings, we included the latest continuous-valued result up to 6 months before the patient’s transfusion. We compiled cumulative bleeding volume over 24 hours prior to transfusion [2]. We visualize cluster characteristics using level 1 and 2 ICD-10 categories and primary procedures and diagnoses compiled in our earlier work [2]. For further details on data compilation, see Supplement Section S1.1.

### 2.2 Ethics

The study followed GDPR, national Act 552/2019 on secondary health and social data use, and was approved by HUS Helsinki University Hospital (HUS/579/2022). The data was pseudonymized by the institution and analysed on a secure analytics platform HUS Acamedic.

### 2.3 Feature selection and data preprocessing

We compiled patient-level data during and prior to the first transfusion episode. Unit-level transfusion data was converted to crosssectional by selecting the first transfusion episode, created by grouping transfusions occurring up to three hours apart. Occasionally missing time annotations were imputed [2]. Each episode was labeled with three binary variables denoting administration of red blood cell (RBC), plasma or platelet transfusion, or their combination. Patients lacking product information were excluded.

Data was split into training (80%), test (20%), and validation (20% of training) sets with balanced labels. To improve stability, diagnosis codes and laboratory values with prevalences below 1% and 70%, respectively, were excluded. Patients without qualifying diagnoses or laboratory values were excluded.

All preprocessing was fit solely on training data to avoid information leakage. Continuous variables were modelled as Gaussian or log-normal (see Supplement Section S1.1), with log-normal variables log-transformed and treated as Gaussian. Outliers (Supplement Equation S1) were set to missing. Missing values were imputed with *IterativeImputer*(Scikit-learn [16]). Binary variables were one-hot encoded. Continuous variables were standardized to zero mean and unit variance instead of min-max scaling to prevent binary features (90% of inputs) dominating latent space formation.

### 2.4 TR-ADE - *Transfusion Recipient Adaptive Deep Embedding*

VAEs are essentially generative models that learn a low-dimensional representation of complex data useful for imputing, denoising, and generating data. Since latent representations of VAEs often approximate manifolds of input data, clustering can be applied externally [17], or incorporated via a GMM prior [18, 19]. To enable clustering on an adjustable level alongside classification of transfused products, we incorporated a GMM-prior VAE with a classification module. A similar approach has been applied to standard-normal VAEs [20]. We extend the model to clustering.

The GMM-prior VAE [18] assumes a generative process, where (*i*) an unobserved cluster label *c*is drawn from a categorical distribution with prior *π*over predefined number of *K*clusters, (*ii*) an unobserved latent variable *z*∈ ℝ^*L*^ is sampled from a Gaussian conditioned on *c*with parameters 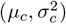, and (*iii*) observed data *x*is generated from *z*via decoder *d*_*θ*_(·), an artificial neural network (ANN) parameterized by *θ*:

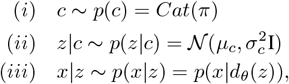

where

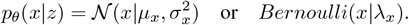

To incorporate simultaneous classification in semisupervised manner, we introduce three observed class labels *y*= {*y*^1^, *y*^2^, *y*^3^}, one for each blood product type. The classifier models the joint distribution over the classification labels:

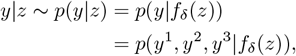

where *f*_*δ*_(*·*) is the classifier parameterized by *δ*.

The true posterior distribution (Supplement Equation S2) is approximated with a surrogate distribution

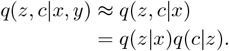

See Supplement Equation S3 for derivation. The first component represents the probabilistic encoder parameterized by *ϕ*used to infer latent variable *z*from observed data *x*(Supplement Equation S4). The second component is computed directly during training (Supplement Equation S5).

The model is trained by maximizing the Evidence Lower-Bound (ELBO) objective, derived from the Kullback-Leibler (KL) divergence between the true and the approximate posterior distributions. The augmented ELBO can be expressed in terms of reconstruction loss, KL-divergence between the approximate posterior and the prior of the latent variables, and classification loss weighted by *γ*:

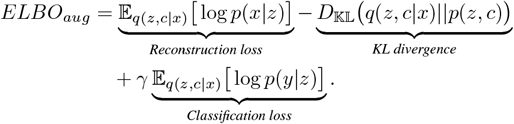

In practice we minimize the negative augmented ELBO. **Figure 1** displays the high-level structure of TR-ADE (see Supplement Figure S1 for details). Here, the ANN parameters (*ϕ, θ, δ*) as well as the GMM prior parameters (*π*, **M**, Σ) are trainable and optimized during training (Supplement Section S1.2.2).

**Figure 1.**
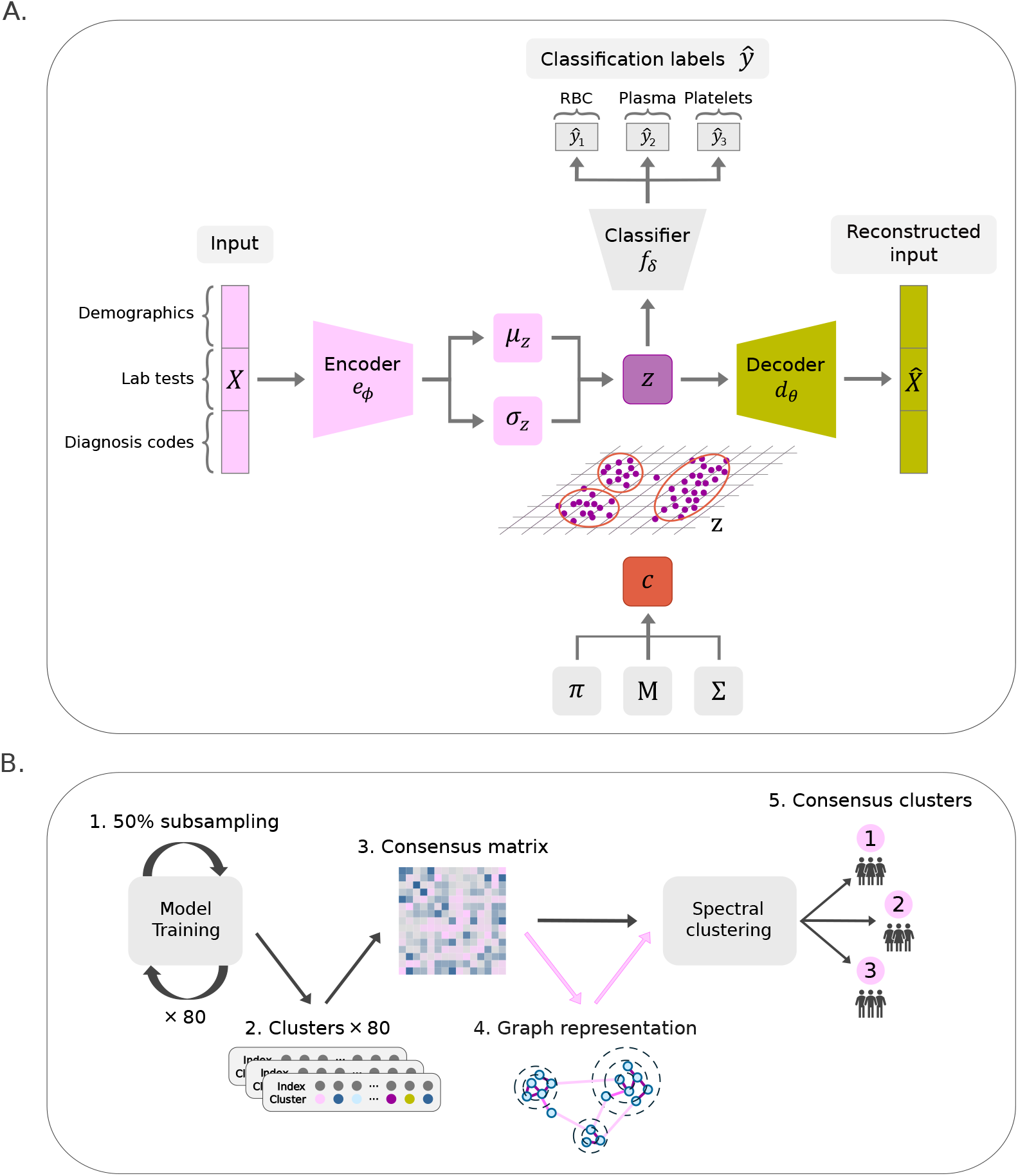
Model architecture and illustration of the clustering pipeline. A) The encoder of TR-ADE generates the parameters of *z*. To shift the stochasticity of the sampling process outside gradient computations during backpropagation, reparameterization trick is applied to get the latent sample that is given as an input to the decoder and the classifier. The parameters of the GMM prior (*π*, **M**, Σ) are optimized simultaneously with the ANN parameters. B) TR-ADE was trained 80 times with subsampled data. For robustness, the resulting clusters were used to construct a consensus matrix which is represented as a graph to determine the number of clusters to be used in spectral clustering.

### 2.5 Clustering procedure

TR-ADE was optimized with Adam [21] and structural hyperparameters (Supplement Table S1) were chosen relative to dataset size to simplify training and limit decoder expressiveness, reducing posterior collapse. Grid search over latent dimension (*L*) and classification weight (*γ*) was evaluated using areas under the receiver operating characteristic curve (AUROC) and the precision-recall curve (PR-AUC), Silhouette Score (Sil-score) [22] and Calinski-Harabasz index (CHI) [23] (Supplement Section S1.3).

TR-ADE assigns hard clusters alongside classification labels but optimizing the number of clusters (*K*) generally requires testing multiple values [24, 25]. In TR-ADE, random seed variations, particularly in parameter initialization, slightly affect clustering, and a large *K*with trainable prior probabilities can leave some clusters unused. To ensure robustness, we applied an ensemble approach using modified consensus clustering [15, 17] (Figure 1). TR-ADE was trained 80 times with different random seeds and 50% subsampling of training data, using the hyperparameters in Supplement Table S2. A consensus matrix [15] (*N*× *N*) of a dataset of size *N*recorded the fraction of runs where patients *i*and *j*were assigned to the same cluster, considering only subsamples including both. We applied spectral clustering [26] (Scikit-learn [16]) to the consensus matrix, with the number of clusters chosen from its graph representation (Supplement Section S1.4).

### 2.6 Cluster evaluation

To identify cluster-enriched features, ICD-10 codes (levels 1 and 2), sex, and binary variable of weekend transfusion were evaluated with *χ*^2^-test (Scipy [27]) and logarithmic odds ratios (logOR) (Supplement Section S1.5). Binary features were included if available for ≥ 20 patients. Continuous variables, including age, laboratory results, blood product distributions, and bleeding were tested with two-sided Mann-Whitney U-test (SciPy [27]) for significant differences in distributions between the cluster and all other data, including variables with ≥ 20 observations per group to ensure statistical power. P-values were corrected for multiple testing using Benjamini-Hochberg with a False Discovery Rate (FDR) of 0.1% (Statsmodels [28]). Variables with adjusted *p <*0.01 were considered significant.

Finally, the robustness of clustering pipeline was evaluated with repeated runs with varying *γ*(Supplement Section S1.6), and the formed clusters were compared against primary diagnoses and procedure categories identified in prior work [2].

## 3 Results

### 3.1 Data description

The final dataset (**Table 1**) included 19,629 unique patients and 153 variables: 135 diagnosis codes, 12 laboratory parameters, sex, age, BMI, and three classification labels. Modified consensus clustering was applied on training data from 15,704 patients, while a test set of 3,925 patients was used to evaluate generalization of the pipeline to unseen data.

**Table 1.** Descriptive characteristics of the complete dataset. Abbreviation IQR: interquartile range.

| Feature | Statistics |  | Completeness (%) |
| --- | --- | --- | --- |
| **Sex, N** | 19628 |  | > 99.9 |
| Female, N | 11122 |  | 56.7 |
| Male, N | 8506 |  | 43.3 |
| **Age at transfusion (years), median (IQR)** | 69.0 | (53.0, 79.0) | 100 |
| **Body mass index ($\text{kg}/\text{m}^2$), median (IQR)** | 25.4 | (22.4, 29.3) | 80.2 |
| **Blood product recipients, N** |  |  |  |
| RBC recipients | 17987 |  | 91.6 |
| Plasma recipients | 2027 |  | 10.3 |
| Platelet recipients | 1991 |  | 10.1 |
| **Laboratory tests, median (IQR)** |  |  |  |
| Hemoglobin (g/l) | 84.0 | (76.0, 101.0) | 95.2 |
| Leukocytes (E9/l) | 8.6 | (6.3, 12.1) | 95.2 |
| Thrombocytes (E9/l) | 220.0 | (156.0, 303.0) | 95.2 |
| Erythrocytes (E12/l) | 2.9 | (2.5, 3.6) | 95.2 |
| Mean corpuscular volume of red cells (MCV) (fl) | 92.0 | (87.0, 97.0) | 95.2 |
| Hematocrit (%) | 26.0 | (24.0, 31.0) | 95.2 |
| Mean hemoglobin mass of red blood cell (MCH) (pg) | 30.0 | (28.0, 32.0) | 95.2 |
| Red cell distribution width (RDW) (%) | 15.0 | (13.0, 16.0) | 95.1 |
| Mean Corpuscular Hemoglobin Concentration (MCHC) (g/l) | 325.0 | (314.0, 335.0) | 95 |
| Creatinine ($\mu\text{mol}/\text{l}$) | 76.0 | (58.0, 106.0) | 90.1 |
| Glomerular filtration rate (ml/min/1.73) | 79.0 | (51.0, 97.0) | 89.7 |
| Thromboplastin time (PT) (%) | 81.0 | (64.0, 98.0) | 75.7 |
| **Main ICD-10 diagnosis categories, N** |  |  |  |
| I00-I99 Diseases of the circulatory system | 9353 |  | 47.6 |
| V01-Y98 External causes of morbidity and mortality | 5111 |  | 26 |
| E00-E90 Endocrine, nutritional and metabolic diseases | 5034 |  | 25.6 |
| K00-K93 Diseases of the digestive system | 4695 |  | 23.9 |
| S00-T98 Injury, poisoning and certain other consequences of external causes | 4304 |  | 21.9 |
| C00-D48 Neoplasms | 4111 |  | 20.9 |
| D50-D89 Diseases of the blood and blood-forming organs and certain disorders involving the immune mechanism | 3579 |  | 18.2 |
| N00-N99 Diseases of the genitourinary system | 3510 |  | 17.9 |
| M00-M99 Diseases of the musculoskeletal system and connective tissue | 2762 |  | 14.1 |
| J00-J99 Diseases of the respiratory system | 2215 |  | 11.3 |
| G00-G99 Diseases of the nervous system | 2002 |  | 10.2 |
| O00-O99 Pregnancy, childbirth and the puerperium | 1701 |  | 8.7 |
| A00-B99 Certain infectious and parasitic diseases | 1566 |  | 8 |
| F00-F90 Mental and behavioural disorders | 1276 |  | 6.5 |
| Z00-ZZB Factors influencing health status and contact with health services | 1193 |  | 6.1 |
| H00-H59 Diseases of the eye and adnexa | 1094 |  | 5.6 |
| L00-L99 Diseases of the skin and subcutaneous tissue | 719 |  | 3.7 |
| U00-U99 Codes for special purposes | 645 |  | 3.3 |
| H60-H95 Diseases of the ear and mastoid process | 234 |  | 1.2 |

### 3.2 Data-driven phenotypes

#### Cluster 1 – Neoplasms & Digestive system

We identified six distinct clusters of transfusion recipients (**Figure 2**) based on hyperparameter configuration determined using grid search (Supplement Section S2.1). Cluster 1 (‘Neoplasms & Digestive system’; N=3,844) was enriched with patients suffering from digestive system diseases (K00-K93) and neoplasms (C00-D48). This cluster likely represents cancer patients and those with gastrointestinal conditions requiring transfusions (Supplement Figure S4). This cluster had slightly less dispersed erythrocyte count (B-Eryt) and hematocrit (HKR) compared to the overall transfusion recipient population (Supplement Figure S5).

**Figure 2.**
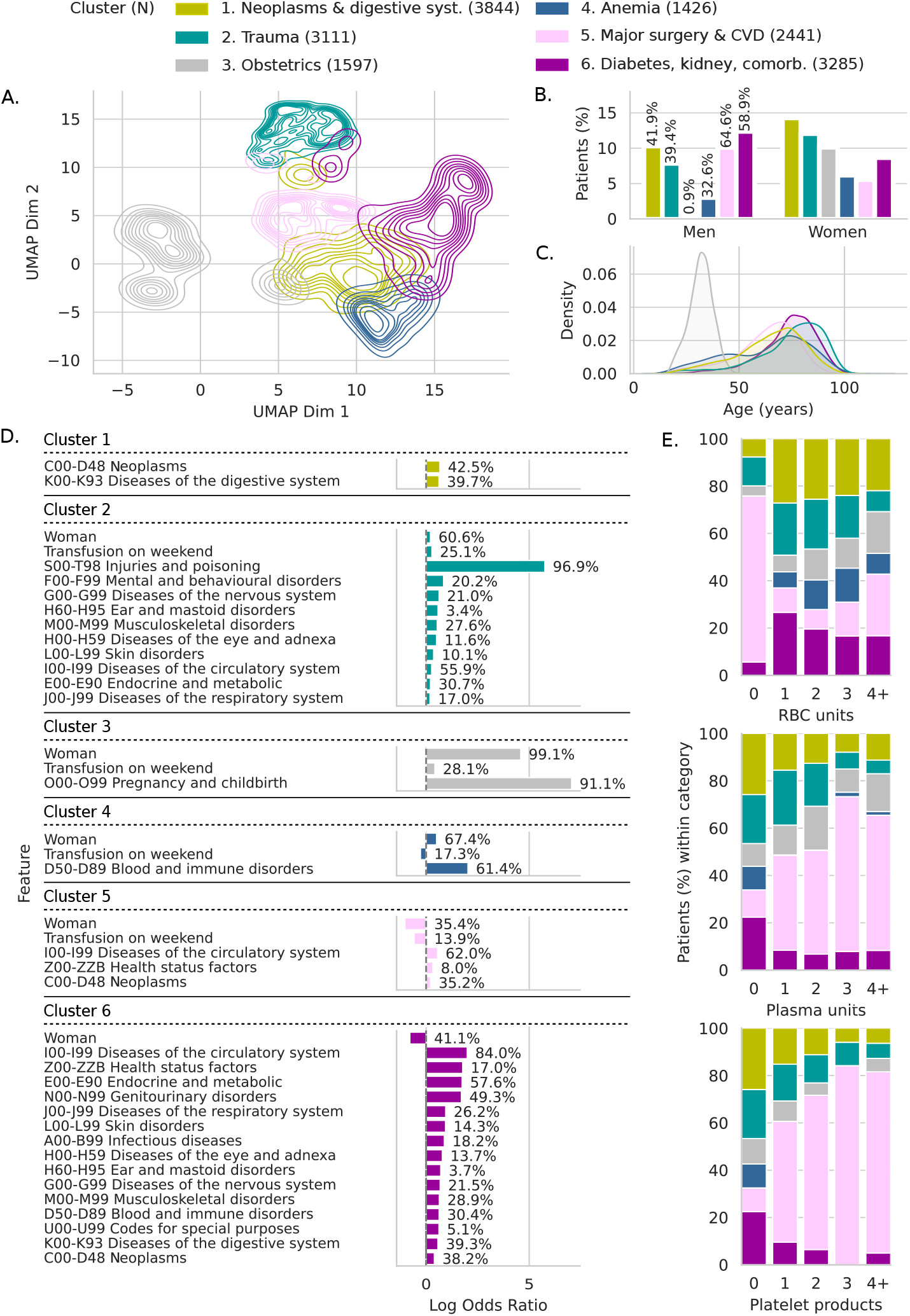
Six distinct data-driven phenotypes of transfusion recipients. A) Example of UMAP [29] projected latent space of TR-ADE trained with hyperparameters in Table S2, patients colored according to consensus clusters, B) sex and, C) age distributions of clusters, D) subset of statistically significant features within clusters, with log-OR and frequency of feature within the cluster, and E) cluster-wise product distributions over transfusion episodes.

#### Cluster 2 – Trauma

Cluster 2 (‘Trauma’; N=3,111) was notable for high prevalence of injury, poisoning and external cause-related diagnoses (S00-T98), i.e. trauma patients (**Figure 2**). Injuries to the hip and thigh (S70-S79) had the highest prevalence and log-OR in this cluster, with 40 other enriched ICD-10 level 2 categories primarily related trauma or diagnosis codes typical to the age group (Supplement Figure S4). Transfusions in these patients took place statistically significantly on weekends. Laboratory results align with an average transfusion recipient (Supplement Figure S5).

#### Cluster 3 – Obstetrics

Cluster 3 (‘Obstetrics’; N=1,597) was distinguished as a unique subgroup of females, with 75% of patients aged 36 or younger (Supplement Table S3). The diagnoses were predominantly related to pregnancy, childbirth, and the puerperium (O00-O99) (**Figure 2**), a well-known subgroup of obstetric transfusion recipients [1]. Laboratory test profiles of this cluster (Supplement Figure S5) are typical to younger, pregnant population, with normal coagulation screening (P-PT, P-APTT), lower ferritin levels indicating iron deficiency, and lower haptoglobin (P-Haptog) levels suggestive of hemolysis related to pregnancy complications. Transfusions took place statistically significantly on weekends (**Figure 2**) and pre-transfusion bleeding was on average the highest in this cluster (**Figure 3**).

**Figure 3.**
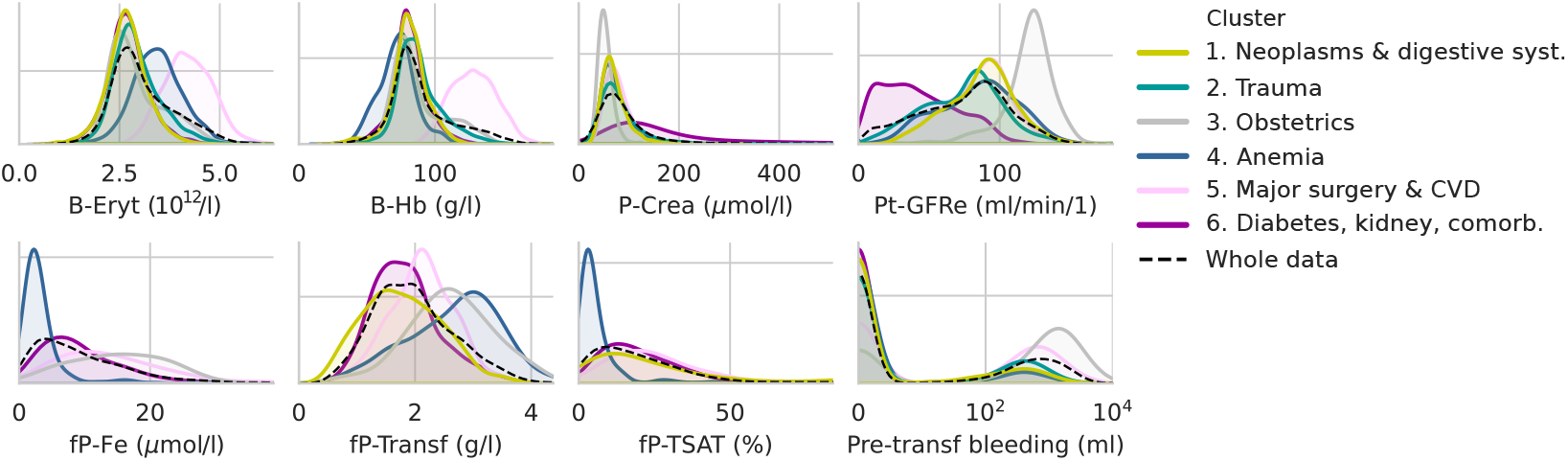
Identified clusters demonstrate unique patterns in laboratory results and bleeding. Sample of statistically significant differences in normalized laboratory variable distributions and bleeding 24 hours prior to transfusion episode.

#### Cluster 4 – Anemia

Cluster 4 (‘Anemia’; N=1,426) had a high proportion of women and more than 60% diagnosed with diseases of the blood and immune system (D50-D89), with highest log-OR and prevalences on nutritional anemias (D50-D53) and aplastic and other anemias (D60-D64). Noninflammatory disorders of female genital track (N80-N98) including abnormal uterine bleeding, and benign neoplasms (D10-D36) are also enriched in this cluster (Supplement Figure S4). Nearly all patients in this cluster received only RBC transfusions (**Figure 2**). Typical findings in red cell indices together with low plasma iron concentration (fP-Fe) and low transferrin saturation (TSAT) indicate iron deficiency anemia (Supplement Figure S5).

#### Cluster 5 – Major surgery and cardiovascular diseases

Cluster 5 (‘Major surgery & CVD’; N=2,441) primarily consisted of male patients, with a high prevalence and log-OR of diseases of the circulatory system (I00-I99) (**Figure 2**), suggesting that many patients underwent cardiovascular surgery or organ transplantation, supported by an increased frequency of plasma and platelet transfusions (**Figure 2**). Of all procedures, these patients most frequently underwent operations associated with transfusion episodes. Furthermore, patients had relatively high HKR and B-Eryt levels (Supplement Figure S5), and on average high pretransfusion bleeding (**Figure 3**).

#### Cluster 6 – Diabetes, kidney disease, other comorbidities

Cluster 6 (‘Diabetes, kidney, comorbidities’; N=3,285) contained a diverse patient population with diagnoses related to circulatory (I00-I99), endocrine (E00-E90), and genitourinary (N00-N99) diseases (**Figure 2**). Enriched diagnoses including glomerular diseases (N00-N08), renal failure (N17-N19), hypertensive diseases (I10-I15), diabetes mellitus (E10-E14) with 55 other ICD-10 level 2 categories (Supplement Figure S4) suggests this cluster captures patients with complex comorbidities and/or requiring dialysis. Laboratory results align with this categorization with elevated creatinine levels and reduced glomerular filtration rate (Pt-GFRe) in this cluster (Supplement Figure S5).

### 3.3 Robustness of clustering compared to primary diagnoses and procedures

Figure 4. displays the association between primary diagnoses, final clusters (N=13,455), and procedures (N=5,962). For all clusters, primary diagnoses align with cluster-enriched diagnoses. Approximately half of ‘Trauma’ patients have undergone a procedure related to musculoskeletal system, whereas only a small fraction of patients in ‘Anemia’ cluster have undergone a procedure associated with transfusion. Most patients in ‘Major surgery & CVD’ have undergone a procedure, including procedures for heart and major thoracic vessels.

Increasing the weight *γ*of product classification on total loss alters clustering primarily in two ways. First, ‘Trauma’ patients increasingly shift to ‘Major surgery & CVD’ cluster (**Figure 4**), reducing the size of ‘Trauma’ cluster. This is evident for primary diagnoses, as patients with injuries and poisoning (S00-T98) map to ‘Trauma’ at *γ*= 0 but mostly to ‘Major surgery & CVD’ at *γ*= 2, with a similar trend for procedures. Second, clusters ‘Neoplasms & Digestive system’ and ‘Diabetes, kidney, comorbidities’ exchange patients across *γ*, with a slight growth of the latter, suggesting these clusters overlap more compared to other clusters. Overlapping was also observed with ICD level 2 categories enriched in multiple clusters (Supplement Figure S6).

**Figure 4.**
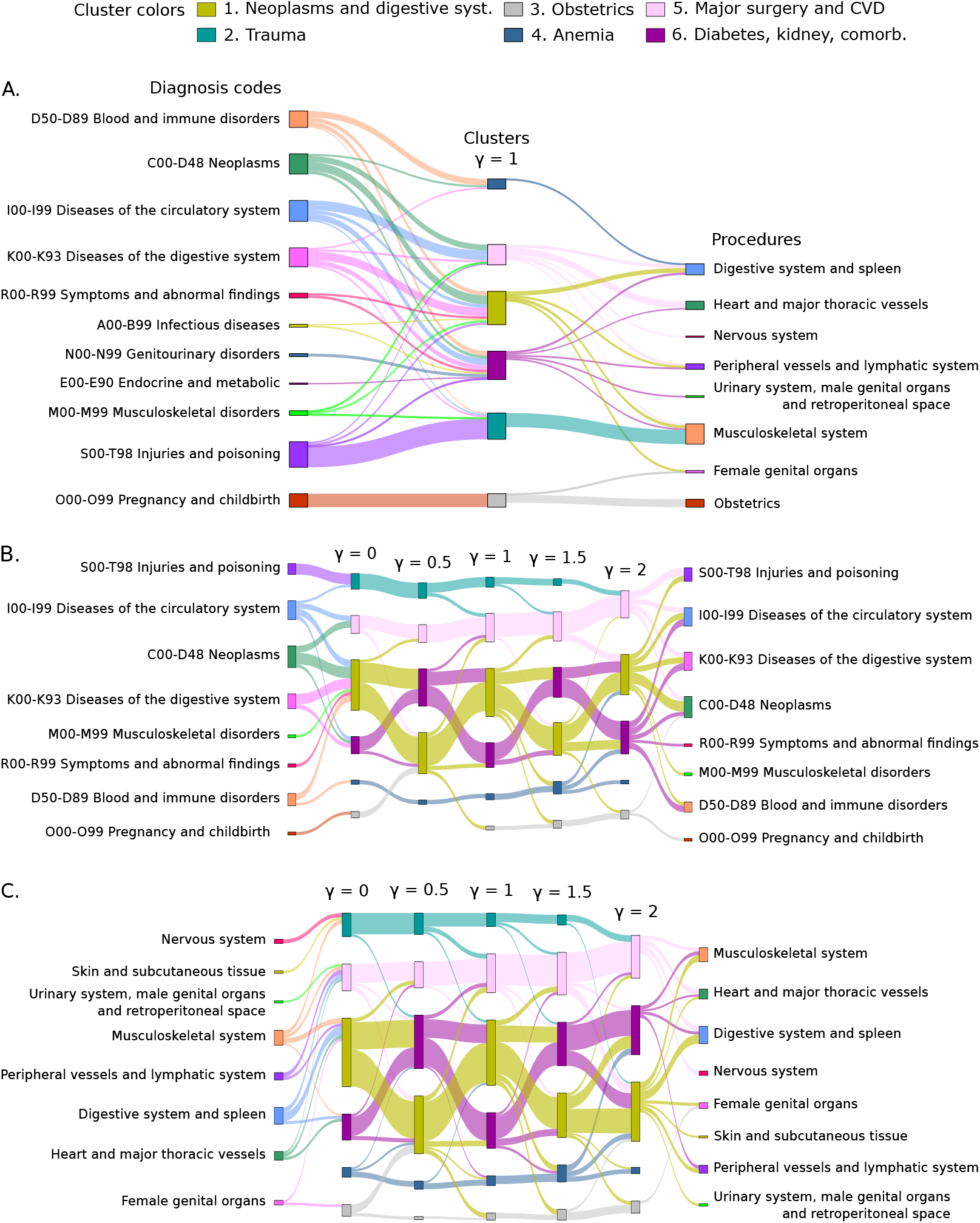
Flow diagrams reveal robustness of clustering and effect of classification weight gamma to cluster composition. A) Primary diagnoses and procedures of transfusion episodes of all patients, excluding flows of *<*100 patients (N=13,455). B) Primary diagnoses of transfusion episodes of mismatched patients excluding flows of *<*40 patients (N=1,888). C) Primary procedures of transfusion episodes of mismatched patients excluding flows of *<*20 patients (N=2,026).

### 3.4 Evolvement of consensus clustering over *γ*

Increasing classification weight *γ*affects consensus clustering in several ways. Larger differences in *γ*between pairwise comparisons yield more mismatched patients (Supplement Figure S7). With higher *γ*, total loss and reconstruction loss rise for training data, while classification loss and KL divergence decrease (Supplement Table S4).

Plasma AUROC remains stable across *γ*(**Figure 5**), while RBC and platelet AUROC increase with *γ*(Supplement Tables S5-S7). However, overfitting appears for *γ >*1. PR-AUC shows similar trends, except plasma declines beyond *γ*= 0.75. Clustering metrics remain stable until *γ*= 1, then deteriorate (Supplement Table S8). Although the AUROC is substantially higher than on a random classifier, PR-AUC better reflects overall performance on this imbalanced dataset, where minority class fractions are within 10 *±*1.6% of the dataset. Overall, classification performance on test data is moderate across *γ*values.

**Figure 5.**
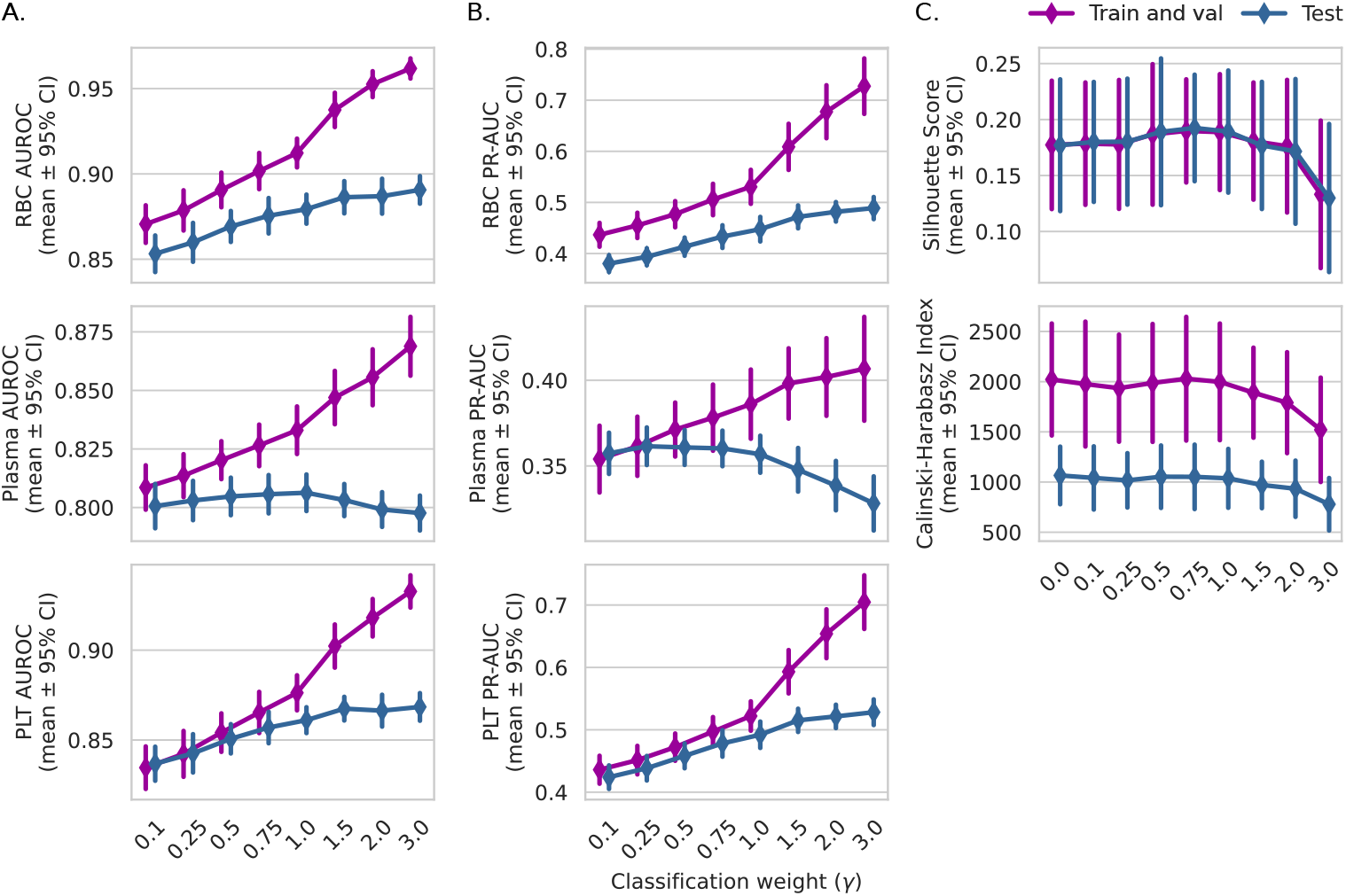
Evolution of consensus clustering metrics over varying classification weight *γ*. A) RBC and platelet AUROC increase with *γ*: RBC from 0.853 *±*0.011 (*γ*= 0.1) to 0.891 *±*0.008 (*γ*= 3); platelets from 0.837 *±*0.009 to 0.868 *±*0.008, and *γ*values *>*1 begin to overfit the model. Plasma classification accuracy is not significantly affected by *γ*values larger than zero, with peak at 0.806 *±*0.008 at *γ*= 1 (See Supplement Tables S5-S7). B) PR-AUC values are 0.472 *±*0.048 and 0.476 *±*0.039 at *γ*= 1 for RBC and platelets, respectively. PR-AUC for plasma (0.364 *±*0.033 at *γ*= 1) declines beyond *γ*= 0.75. Calculated for minority class. C) Clustering metrics. Sil-score ranges from 0.177 (*γ*= 0) to 0.189 (*γ*= 1), while CHI decreases slightly; from 2021 to 1998 (training set) and from 1066 to 1037 (test set) (Supplement Table S8).

## 4 Discussion

We characterized a population of adult transfusion recipients in a data-driven manner using TR-ADE, a VAE with a novel combination of a GMM prior and a classification component. Our findings refine the categorization of transfusion recipients recognized in previous literature [3–6, 8–10], but extend the phenotypes over diagnosis history and multidimensional patient feature profiles. The method demonstrates robust patient grouping with respect to primary diagnoses and procedures, while simultaneously classifying the transfused blood product.

Our findings highlight the heterogeneity of transfusion recipients and, instead of relying on human-driven categorization and population level statistics on key features, we show that the phenotypes are characterized by a wider variety of attributes and their associations. Thus, this study presents a data-driven, integrative view of a transfusion recipient. Both ‘Obstetrics’ and ‘Trauma’ clusters are associated with weekend transfusions, which aligns with the randomness of both events. Although anemia predicts transfusion in multiple patient groups [30], the ‘Anemia’ cluster seems to capture those with chronic anemia due to either a hematologic disease or slow bleeding for non-malignant causes (e.g. gynecologic bleeds).

Bleeding in ‘Major surgery & CVD’ suggests hemorrhage in surgery, rather than a chronic condition, necessitating transfusion. These results align with the common knowledge that major surgeries often require transfusions, a finding supported also by our previous work [2]. Impaired renal function was typical in the ‘Diabetes, kidney, comorbidities’ cluster, implying those with complex comorbidities or on dialysis form a distinct patient group receiving transfusions.

While each cluster is characterized by a distinct set of enriched features, we observed some overlap between ‘Neoplasms & Digestive system’ and ‘Diabetes, kidney, comorbidities’ clusters. These patients are likely multimorbid, sharing characteristics of both clusters, and assignment into clusters depends on the model assumptions. However, patients traversing between the clusters when the classification weight is adjusted likely lack distinct features for a separate group within our data and model.

Our approach captures the heterogeneity of the population, but also the heterogeneity in blood demand. Transfusion decisions made by clinicians are affected by various patient-related features, such as a combination of diagnoses, planned procedures, and laboratory values, observable to the decision-maker. Phenotype-informed grouping can clarify the structure of the demand drivers, possibly enabling the implementation of simpler yet effective policies and systems across the blood supply chain.

Despite the robustness of the presented clusters, this study has limitations. Choices in planning and execution of data validation, compilation, matching, exclusion criteria, feature engineering, or preprocessing directly affect our data, and thus model quality. Although fine-grained, the used EHR data may contain systematic errors [2]. Furthermore, consensus clustering and grid search illustrate that combining unsupervised clustering with discriminative classification results in a trade-off between clustering coherence and classification accuracy. Classification weight *γ*adjusts how class information influences the latent space, and modifying it shifts some patients between clusters, making the clustering non-exclusive. Finally, the pipeline generalizability has not been evaluated on data from another EHR system.

The strengths of this study include comprehensive high-resolution data, a robust clustering pipeline, and clinically relevant results coupled with model explainability. Clusterings remain stable on ≥87% of data across *γ*, while patients shifting clusters provide insights on how the classification module shapes the latent space. Furthermore, although transfusion populations vary with country income level and patient demographics [1], and differences in healthcare systems may further limit the generalizability of transfusion population studies, our selection of methods ensure relevancy in comparable settings.

## 5 Conclusions

Data-driven characterization of transfusion recipients provides a previously unexplored view on the multidimensional phenotypes of patients requiring blood transfusion. The developed model, based on novel methodology, helps to understand the heterogenous nature of transfusion recipients and provides a basis for future attempts to utilize EHR data to optimize blood supply chain operations and transfusion management practices.

## Data Availability

EU and national legislation restrict the distribution of personal health data. No data is made available outside authorized use.

## Funding statement

The study was supported by The Research Fund of the Finnish Red Cross Blood Service.

## Data and code availability statement

EU and national legislation restrict the distribution of personal health data.

The source code for TR-ADE has been implemented using Keras 3 [31] with Tensorflow 2.17 [32] backend and is available in Github.

## CRediT authorship contribution statement

**Elissa Peltola:** Writing - original draft, Writing – review & editing, Software, Methodology, Investigation, Formal Analysis, Data curation, Validation, Visualization, Conceptualization. **Esa Turkulainen:** Writing – review & editing, Data curation, Funding acquisition. **Markus Heinonen:** Writing – review & editing. **Mikko Arvas:** Writing – review & editing, Funding acquisition. **Minna Ilmakunnas:** Writing – review & editing, Project administration, Funding acquisition, Validation. **Miika Koskinen:** Writing – review & editing, Methodology, Conceptualization, Funding acquisition, Project administration, Supervision, Validation.

## Declaration of competing interest

The authors declare no competing interests.

## Notes

### Competing Interest Statement

The authors have declared no competing interest.

### Author Declarations

In accordance with Finnish legislation (Act on the Secondary Use of Health and Social Data 552/2019), ethical committee approval or informed consent is not required for retrospective registry studies. This study complied with the General Data Protection Regulation (GDPR) and received approval from HUS Helsinki University Hospital (HUS/579/2022, 17 October 2022).

