## Supplementary Material for "Deep latent variable modelling reveals clinically significant subgroups among transfusion recipients"

### ONLINE SUPPLEMENTARY MATERIAL FOR:
### DEEP LATENT VARIABLE MODELLING REVEALS CLINICALLY SIGNIFICANT SUBGROUPS AMONG TRANSFUSION RECIPIENTS

**Elissa Peltola**[1,*] **Esa Turkulainen**[2] **Markus Heinonen**[3] **Mikko Arvas**[2] **Minna Ilmakunnas**[4, 2]

**Miika Koskinen**[5]

October 29, 2025

[1]IT Management, HUS Helsinki University Hospital
[2]Research and Development, Finnish Red Cross Blood Service
[3]Department of Computer Science, Aalto University
[4]Department of Anesthesiology and Intensive Care Medicine, HUS Helsinki University Hospital and University of Helsinki
[5]Diagnostic Center & New Children's Hospital, HUS Helsinki University Hospital and University of Helsinki

**S3 Experimental research** **18**

### S1 Supplement methods

#### S1.1 Data and preprocessing details

Body mass index (BMI) was either directly available or computed using available height and weight with equation

$$BMI = \frac{weight(kg)}{height(m)^2}.$$

Missing weight measurements were imputed as the previous non-missing value with one-month tolerance, and height unambiguously as the last non-missing value before the start of transfusion episode. Laboratory tests P-Ca-Ion and P-Ca-IonA, as well as S-Ca-Ion and S-Ca-IonA were unified to P-Ca-Ion and S-Ca-Ion by taking the average of these values, if both were present.

Likelihoods of continuous variables were determined based on smaller Akaike Information Criterion (AIC) for fitted normal and log-normal distributions. However, if absolute skewness exceeded 1, the variable was set to log-normal. The outliers of continuous variables were defined by values outside

$$\big[Q_1 - 1.5 \times IQR,\ Q_3 + 1.5 \times IQR\big] \tag{S1}$$

where $Q_1$ and $Q_3$ are 0.25 and 0.75 quantiles, respectively, and $IQR = Q_3 - Q_1$ is the interquartile range.

#### S1.2 Model derivation and training details

##### S1.2.1 Introduction to deep latent variable modelling

Latent variable models (LVMs) provide a probabilistic framework for capturing hidden structures in potentially high-dimensional data by introducing unobserved latent variables that explain the observed data. These latent variables can be used in numerous tasks like dimension reduction, data generation, imputation, clustering and classification. However, computing the posterior distribution of these latent variables is often intractable due to hidden nature of latent variables and complexity of ground truth distributions. Variational inference (VI) addresses this by approximating the true posterior with a distribution from a tractable distribution family, optimizing its parameters by minimizing the Kullback-Leibler (KL) divergence between the two distributions. This enables efficient approximate inference and representation learning.

Variational Autoencoder (VAE) [1] is a generative model that extends VI to deep learning by combining artificial neural networks (ANN) with probabilistic posterior inference. In VAEs, the encoder learns a variational posterior $q(\cdot)$ that approximates the true posterior $p(\cdot)$ while the decoder reconstructs data from sampled latent variables. The training objective maximizes evidence lower bound (ELBO), derived from the KL divergence of VI, to balance reconstruction accuracy with a regularization term to ensure structured latent space representations. The Vanilla-VAE [1] assumes standard normal prior on latent variables and diagonal Gaussian as the posterior distribution, but the framework can be easily extended to other settings. Here we adapt Gaussian Mixture Model (GMM) prior in the fashion of [2], to encourage clustering of the latent space. Furthermore, we extend the GMM prior VAE model with a classifier module to perform simultaneous classification on transfusion outcome and essentially, to influence the latent space cluster formation.

##### S1.2.2 TR-ADE - Full derivation

Recall the generative process of the GMM prior VAE backbone and the incorporated classification label (Section 2.3 in Original article) of TR-ADE. The joint probability distribution of the observed data and unobserved latent variables is defined as

$$p(x, y, z, c) = p(x|x)p(y|z)p(z|c)p(c).$$

This factorization is essentially derived from the assumed generative process of the observed data. The true posterior of the latent variables can then be derived using Bayes' Theorem:

$$\begin{aligned} p(z, c|x, y) &= \frac{p(x, y, z, c)}{p(x, y)} \\ &= \frac{p(x|x)p(y|z)p(z|c)p(c)}{p(x, y)}. \end{aligned} \tag{S2}$$

This true posterior is approximated with a surrogate that factorizes in the following way:

$$\begin{aligned} p_\theta(z, c|x, y) &\approx q_\phi(z, c|x) \\ &= q_\phi(z|x)q_\phi(c|x, z) \\ &== q_\phi(z|x)q_\phi(c|z). \end{aligned} \tag{S3}$$

The first component of the factorization retains the approach of Vanilla-VAE, as we model the distributions of $z$, i.e. the Gaussian components of the mixture model as diagonal Gaussian distribution, assuming mean-field factorization:

$$\begin{aligned} z|x &\sim q_\phi(z|x) \\ &= q(z|e_\phi(x)) \\ &= \mathcal{N}(\mu_z, \sigma_z^2 I), \end{aligned} \tag{S4}$$

where $e_\phi(\cdot)$ is the encoder, an ANN parameterized by $\phi$. The second component is computed analytically using Bayes' rule:

$$q(c|z) \approx p(c|z) = \frac{p(z|c)p(c)}{\sum_{k=1}^{K} p(z|c=k)p(c=k)}, \tag{S5}$$

where $p(c)$ are the prior probabilities and $p(z|c)$ is the likelihood of drawn sample $z$ belonging to cluster $c$. In addition to utilizing this formulation in soft clustering during loss calculations, it is also used when computing hard cluster assignments for each patient, by selecting such cluster that maximizes the posterior probability $p(c|z)$.

As with the VI framework, the objective function can be derived from minimizing the KL divergence between true and approximate posterior distributions. For the sake of clarity, the parameter subscripts of distributions have been discarded in the following derivations. Recall the true posterior (Equation S2) and approximate posterior (Equation S3). From here we can derive the augmented ELBO objective using the definition of KL divergence:

$$\begin{aligned} D_{\mathbb{KL}}\big(q(z, c|x)||p(z, c|x, y)\big) &= \int q(z, c|x) \log\left(\frac{q(z, c|x)}{p(z, c|x, y)}\right) \mathrm{d}z \\ &= \int q(z, c|x) \log\left(q(z, c|x) - \left(\frac{p(x|z)p(y|z)p(z|c)p(c)}{p(x, y)}\right)\right) \mathrm{d}z \\ &= \mathbb{E}_{q(z,c|x)}\big[\log q(z, c|x)\big] - \mathbb{E}_{q(z,c|x)}\left[\log\left(\frac{p(x|z)p(y|z)p(z|c)p(c)}{p(x, y)}\right)\right] \\ &= \mathbb{E}_{q(z,c|x)}\big[\log q(z, c|x) - \log p(x|z) - \log p(y|z) - logp(z|c) - \log p(c)\big] + \mathbb{E}_{q(z,c|x)}\big[\log p(x, y)\big] \\ &= \mathbb{L}(\phi, \theta, \delta|\mathcal{D}) + \log p(x, y), \end{aligned}$$

where $\mathcal{D}$ is the data $p(x, y)$. The evidence, i.e. marginal log-likelihood of the data $\log p(x, y)$ does not depend on the latent variables and, inherently, the variational parameters we seek in the VI framework to minimize the KL divergence, and thus their expectation w.r.t. $q(z, c|x)$ is the constant itself and can be left out of optimization. Due to the nonnegativity of KL divergence, we get the lower bound for the evidence we seek to maximize:

$$\begin{aligned} \mathbb{L}(\phi, \theta, \delta|\mathcal{D}) + \log p(x, y) &\geq 0 \\ \log p(x, y) &\geq -\mathbb{L}(\phi, \theta, \delta|\mathcal{D}) = ELBO_{aug}. \end{aligned}$$

The augmented ELBO can be written in terms of reconstruction loss, KL divergence between approximate posterior distribution and the prior of the latent variables, and the classification loss:

$$\begin{aligned} ELBO_{aug} &= \mathbb{E}_{q(z,c|x)}\big[\log p(x|z) + \log p(y|z) + \log p(z|c) + \log p(c) - \log q(z, c|x)\big] \\ &= \mathbb{E}_{q(z,c|x)}\big[\log p(x|z)\big] + \mathbb{E}_{q(z,c|x)}\big[\log p(y|z)\big] - \mathbb{E}_{q(z,c|x)}\big[\log q(z, c|x) - \log p(z, c)\big] \\ &= \mathbb{E}_{q(z,c|x)}\big[\log p(x|z)\big] + \mathbb{E}_{q(z,c|x)}\big[\log p(y|z)\big] - \mathbb{E}_{q(z,c|x)}\left[\log \frac{q(z, c|x)}{p(z, c)}\right] \\ &= \mathbb{E}_{q(z,c|x)}\big[\log p(x|z)\big] + \mathbb{E}_{q(z,c|x)}\big[\log p(y|z)\big] - \sum_c \int q(z, c|x) \log \frac{q(z, c|x)}{p(z, c)} \mathrm{d}z \\ &= \mathbb{E}_{q(z,c|x)}\big[\log p(x|z)\big] + \mathbb{E}_{q(z,c|x)}\big[\log p(y|z)\big] - D_{\mathbb{KL}}\big(q(z, c|x)||p(z, c)\big). \end{aligned}$$

By adding the classification weight parameter $\gamma$, we recover Equation 1 in the Original article.

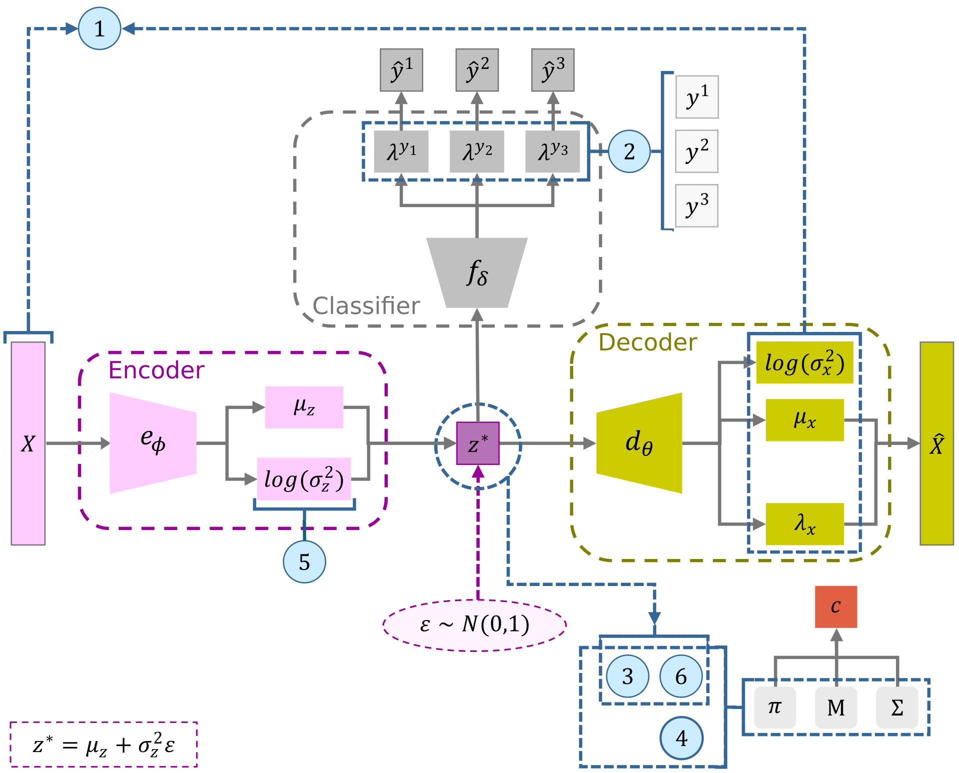

**Figure S1**: Detailed TR-ADE architecture. Terms 1-6 refer to loss terms, i.e. reconstruction loss, classification loss, clustering loss, prior loss, and entropies of the terms of the variational posterior, respectively. The figure displays respective parameters that are used in calculating the terms during training.

We can further express the loss in six components that account for reconstruction, classification and clustering during training:

$$\begin{aligned}
\ell(\phi, \theta, \delta|\mathcal{D}) &= \mathbb{E}_{q(z,c|x)}\big[\log p(x|z) + \log p(y|z) + \log p(z|c) + \log p(c) - \log q(z,c|x)\big] \\
&= \underbrace{\mathbb{E}_{q(z,c|x)}\big[\log p(x|z)\big]}_{1} + \underbrace{\mathbb{E}_{q(z,c|x)}\big[\log p(y|z)\big]}_{2} + \underbrace{\mathbb{E}_{q(z,c|x)}\big[\log p(z|c)\big]}_{3} \\
&\quad + \underbrace{\mathbb{E}_{q(z,c|x)}\big[\log p(c)\big]}_{4} - \underbrace{\mathbb{E}_{q(z,c|x)}\big[\log q(z|x)\big]}_{5} - \underbrace{\mathbb{E}_{q(z,c|x)}\big[\log q(c|z)\big]}_{6}.
\end{aligned} \tag{S6}$$

Figure **S1** gives a detailed depiction of TR-ADE, and how different components are related to calculating the six loss terms.

In the following, we show how the terms 1-6 in Equation S6 are computed in practice.

1. **Reconstruction loss**:
   Let $B$ be the size of minibatch, $I$ be the index set of continuous variables and $J$ be the index set of binary variables. Then,

$$\begin{aligned}
\mathbb{E}_{q(z|x)p(c|z)}\big[\log p(x|z)\big] &= \mathbb{E}_{q(z|x)}\Big[\mathbb{E}_{p(c|z)}\big[\log p(x|z)\big]\Big] & * \\
&= \mathbb{E}_{q(z|x)}\big[\log p(x|z)\big] & \dagger \\
&\approx \frac{1}{B}\sum_{b=1}^{B}\big(\log p_I(x_b|z_b) + \log p_J(x_b|z_b)\big),
\end{aligned} \tag{S7}$$

*Linearity of expectation.
†$p(x|z)$ is constant w.r.t. $p(c|z)$.

where

$$\log p_I(x_b|z_b) = -\frac{1}{2}\sum_{i\in I}\left(\frac{(x_{b,i}-\mu_{b,i})^2}{\sigma^2_{b,i}} + \log\sigma^2_{b,i}\right)$$

and

$$\log p_J(x_b|z_b) = \sum_{j\in J}\big(x_{b,j}\log\mu_{b,j} + (1-x_{b,j})\log(1-\mu_{b,j})\big),$$

that correspond to log-likelihood of Gaussian distribution up to constant factors and log-likelihood of Bernoulli distribution, respectively. To prevent imputation affecting significantly to the reconstruction loss, it was calculated only over observed values.

2. **Classification loss:**

$$\begin{aligned}\mathbb{E}_{q(z|x)p(c|z)}\big[\log p(y|z)\big] &= \mathbb{E}_{q(z|x)}\Big[\mathbb{E}_{p(c|z)}\big[\log p(y|z)\big]\Big] \\ &= \mathbb{E}_{q(z|x)}\big[\log p(y|z)\big] \\ &= \mathbb{E}_{q(z|x)}\big[\log p(y^1,y^2,y^3|z)\big] \\ &\approx \frac{1}{B}\sum_{b=1}^{B}\sum_{j=1}^{3}\log p(y^j_b|z_b),\end{aligned} \tag{S8}$$

where

$$\log p(y^j_b|z_b) = \sum_{k=1}^{K_j} y^j_{b,k}\log(\lambda^{y_j}_{b,k}),$$

and $K_j$ is the number of distinct classes for label $j$, $y^j_{b,k}$ is the $k$th element in one-hot encoded true class label $j$ of data point $b$ in a minibatch, and $\lambda^{y_j}_{b,k}$ is the corresponding predicted label probability.

3. **Clustering loss:**

$$\begin{aligned}\mathbb{E}_{q(z|x)p(c|z)}\big[\log p(z|c)\big] &= \mathbb{E}_{q(z|x)}\Big[\mathbb{E}_{p(c|z)}\big[\log p(z|c)\big]\Big] \\ &= \mathbb{E}_{q(z|x)}\Big[\sum_{k=1}^{K}p(c=k|z)\log p(z|c=k)\Big] \quad {}^{\ddagger} \\ &\approx \frac{1}{B}\sum_{b=1}^{B}\sum_{k=1}^{K}p(c=k|z_b)\log p(z_b|c=k),\end{aligned} \tag{S9}$$

where $p(c|z)$ is calculated according to Equation S5 and $p(z|c)$ as likelihood of sample $z$ belonging to the Gaussian clusters.

4. **Prior loss:**

$$\begin{aligned}\mathbb{E}_{q(z|x)p(c|z)}\big[\log p(c)\big] &= \mathbb{E}_{q(z|x)}\Big[\mathbb{E}_{p(c|z)}\big[\log p(c)\big]\Big] \\ &= \mathbb{E}_{q(z|x)}\Big[\sum_{k=1}^{K}p(c=k|z)\log p(c)\Big] \\ &\approx \frac{1}{B}\sum_{b=1}^{B}\sum_{k=1}^{K}p(c=k|z_b)\log p(c=k),\end{aligned} \tag{S10}$$

where $p(c)$ are the prior probabilities.

5. **Variational posterior loss 1:**

$$\begin{aligned}-\mathbb{E}_{q(z|x)p(c|z)}\big[\log q(z|x)\big] &= -\mathbb{E}_{q(z|x)}\Big[\mathbb{E}_{p(c|z)}\big[\log q(z|x)\big]\Big] \\ &= -\mathbb{E}_{q(z|x)}\big[\log q(z|x)\big] \quad {}^{\S} \\ &\approx \frac{1}{B}\sum_{b=1}^{B}\Big(\frac{1}{2}\log\Big((2\pi e)^L + \det(\Sigma_{z_b})\Big)\Big),\end{aligned} \tag{S11}$$

[‡]Expectation of a categorical variable.

where $\det(\Sigma_{z_b}) = \prod_{l=1}^{L} \sigma^2_{z_b,l}$ as $q(z|x)$ is a diagonal Gaussian and thus we can write

$$-\mathbb{E}_{q(z|x)p(c|z)}\big[\log q(z|x)\big] \approx \frac{1}{B}\sum_{b=1}^{B}\Big(\frac{L}{2}\big(\log(2\pi)+1\big)+\frac{1}{2}\sum_{l=1}^{L}\log\sigma^2_{z_b,l}\Big).$$

6. **Variational posterior loss 2:**

$$\begin{aligned}
-\mathbb{E}_{q(z|x)p(c|z)}\big[\log p(c|z)\big] &= \mathbb{E}_{q(z|x)}\Big[-\mathbb{E}_{p(c|z)}\big[\log p(c|z)\big]\Big] \qquad \text{(S12)}\\
&= \mathbb{E}_{q(z|x)}\Big[\sum_{k=1}^{K} p(c=k|z)\log p(c=k|z)\Big] \qquad \P\\
&\approx \frac{1}{B}\sum_{b=1}^{B}\sum_{k=1}^{K} p(c=k|z_b)\log p(c=k|z_b).
\end{aligned}$$

In practice, the optimization is done by minimizing the negative augmented ELBO rather than maximizing the augmented ELBO. Thus, the negative log-likelihood of Bernoulli distribution in calculating term 1 (Equation S7) becomes binary cross-entropy, and the negative log-likelihood of the categorical distribution in calculating term 2 (Equation S8) becomes cross-entropy loss. Additionally, note that expectations of continuous variables are approximated with mean, where as for the categorical cluster label the expectation is tractable and can be computed exactly. Utilizing this formulation, hard cluster assignments for data points are computed as the cluster that maximizes the posterior probability of Equation S5.

#### S1.3 Training and optimization details

##### S1.3.1 Hyperparameters

The source code for TR-ADE has been implemented using Keras 3 [3] with Tensorflow 2.17 [4] backend. The encoder of TR-ADE has three fully connected hidden layers with 64, 32 and 16 neurons, respectively, with ReLU activations, and two output layers with $L$ neurons, one for each parameter of $z$. The decoder mirrors the encoder with three outputs: two for Gaussian variables and one for binary variables with sigmoid activation. The classifier has one hidden layer with 16 neurons with ReLU activation and three output layers with softmax activations, to extend the model to multiclass setting. All ANN parameters ($\phi$, $\theta$, $\delta$) were initialized with Glorot uniform, GMM prior parameters $\mathbf{M}$ and $\mathbf{\Sigma}$ with Glorot normal and $\pi$ with uniform distribution.

Optimization was performed with Adam [5], a stochastic gradient descent optimization method, by minimizing negative ELBO. We used decaying learning rate with factor of 0.5, minimum learning rate of $10^{-6}$ and patience of 10. Early stopping was used with patience of 50 iterations and minimum change of 0.001 to be considered improvement. Batch size was set to 100, with maximum epochs of 5,000. Both the early stopping and the decaying learning rate monitored the validation loss. For complete list of hyperparameters, see Table **S1**.

##### S1.3.2 Grid search

We evaluated the effect of the latent dimension ($L$) and the classification loss weight ($\gamma$) on optimization, classification, and clustering via grid search with Monte Carlo cross-validation (80/20 split) across five random seeds per parameter combination (Table **S2**), keeping other hyperparameters fixed. For classification, we averaged areas under the receiver operating characteristic curve (AUROC) and the precision-recall curve (PR-AUC); for clustering, we used Silhouette Score (Sil-score) [6] and Calinski-Harabasz index (CHI) [7]. Sil-score measures point similarity to its assigned cluster versus others, while CHI is the ratio of between-cluster to within-cluster dispersion, adjusted for cluster number and sample size. Higher values indicate compact and well-separated clusters.

[§] Definition of differential entropy.

[¶] Entropy of categorical variable.

**Table S1**: Hyperparameters of TR-ADE. *Decoder is a mirror image of the encoder.

| Parameter | Value |
|---|---|
| $L$ (latent dimension) | 3 |
| $K$ (number of clusters) | 15 |
| $\gamma$ (classification weight) | 1 |
| Number of variables to classify | 3 |
| Number of distinct classes for $y_i$ | 2 |
| Encoder hidden layers* | 3 |
| Neurons on encoder hidden layers* | 64, 32, 16 |
| Classifier hidden layers | 1 |
| Neurons on classifier hidden layers | 16 |
| Activation on hidden layers | ReLU |
| Activation on classifier output layer | softmax |
| NN parameter initialization | Glorot uniform |
| GMM parameter $(M, \Sigma)$ initialization | Glorot normal |
| GMM prior $(\pi)$ initialization | Uniform |
| Batch size | 100 |
| NMaximum number of epochs | 5000 |
| Fraction of validation set | 0.2 |
| Setting $\pi$ as trainable parameter | True |
| Optimizer | Adam [5] |
| Decaying learning rate | Keras: ReduceLROnPlateau |
| Factor of decaying learning rate | 0.5 |
| Minimum learning rate | $10^{-6}$ |
| Learning rate patience | 10 |
| Early stopping | Keras: EarlyStopping |
| Early stopping patience | 50 |
| Loss to monitor | 'val_total_loss' |
| Minimum change to be considered improvement | 0.001 |
| Restore best weights | True |
| Direction of improvement | 'min' |

**Table S2**: Hyperparameter combinations over grid search and consensus clustering.

| | Hyperparameter | |
|---|---|---|
| | $L$ | $\gamma$ |
| Grid search | 2, 3, 5 | 0, 0.1, 0.5, 1.0, 1.5, 2.0, 3.0 |
| Consensus clustering | 3 | 0, 0.1, 0.25, 0.5, 0.75 1.0, 1.5, 2.0, 3.0 |

##### S1.4 Determining number of clusters in modified consensus clustering

To determine the number of clusters in advance as required in spectral clustering, we constructed a graph representation of the subsampled consensus matrix $(n = 9000)$ by treating patients as nodes and values of the consensus matrix as edge weights, with larger values indicating that the patients are closer to each other. The graph representation was constructed using NetworkX [8] and *spring_layout* algorithm, which applies Fruchterman-Reingold force-directed

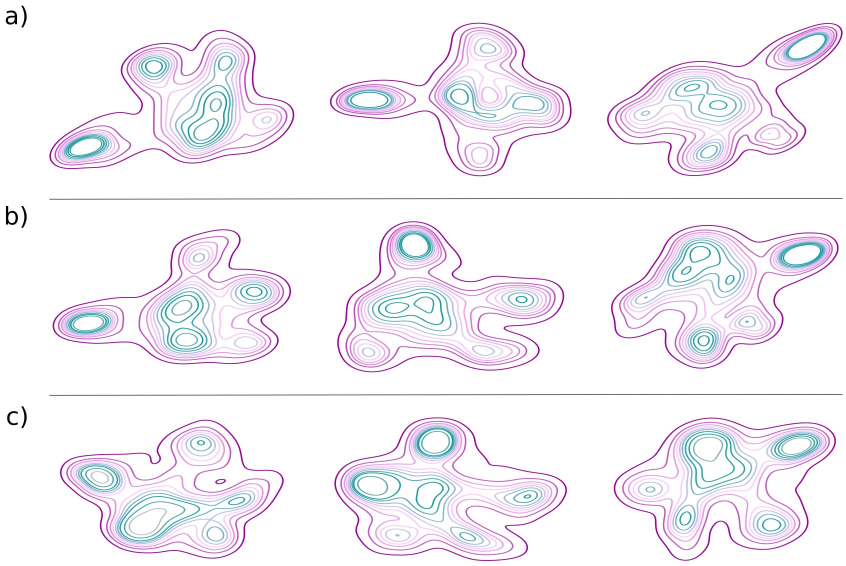

**Figure S2**: Density plots of graph representation of consensus matrix for a) $\gamma = 0$, b) $\gamma = 1$ (used in final analysis), c) $\gamma = 2$.

layout [9] to minimize the energy of the system, where nodes are modelled as repellent charges and edges as springs pulling nodes together with a force proportional to the edge weight.

Process was repeated with different random seeds for classification weights $\gamma \in \{0, 1, 2\}$. By visual inspection (Figure **S2**) of the density plots of the graph representations, we selected the number of clusters to be six.

##### S1.5 Odds ratio

Odds ratio (OR) is a ratio between the empirical odds of a sample belonging to the evaluated cluster compared to the empirical odds of a sample belonging to the rest of the data:

$$\mathrm{OR}(f, c)) = \frac{O_1(f, c)}{O_2(f, c)}, \tag{S13}$$

for feature $f$ and cluster $c$, where

$$O_1(f, c) = \frac{\text{\# patients in cluster } c \text{ with feature } f}{\text{\# patients in cluster } c \text{ without feature } f}, \tag{S14}$$

and

$$O_1(f, c) = \frac{\text{\# patients not in cluster } c \text{ with feature } f}{\text{\# patients not in cluster } c \text{ without feature } f}. \tag{S15}$$

##### S1.6 Robustness of clustering with respect to the weight parameter $\gamma$, primary diagnoses and procedures

While TR-ADE combines VAE with a discriminative classifier, we evaluated how latent representation, classification accuracy, and clustering results varied with the weight parameter $\gamma$. The clustering pipeline was repeated across $\gamma$ values (Table S2), keeping other hyperparameters fixed (Table S1). Spectral clustering used six clusters, chosen from a graph representation at $\gamma = 1$, $L = 3$. Performance on training and test sets was evaluated over 80 runs using AUROC, PR-AUC, Sil-score, CHI and positive predictive value (PPV).

To assess how final clusters align with primary ICD diagnoses and procedure categories identified in prior work [10], Sankey diagrams were drawn from repeated consensus clusterings, excluding flows with $< 100$ patients for clarity. Clustering results across $\gamma$ values were compared pairwise by labeling the six clusters by feature profiles. To examine changes with increasing $\gamma$, we compared consecutive clusterings and identified mismatches, defined as patients assigned to different clusters across compared solutions. We included patients with at least one mismatch in the Sankey diagrams, showing cluster evolution across increasing $\gamma$ relative to primary diagnoses and procedures. Flows with $< 40$ and $< 20$ patients for diagnoses and procedures, respectively, were omitted.

### S2 Supplement results

#### S2.1 Grid search

Grid search showed AUROC and PR-AUC generally improved with larger latent dimension $L$ and classification weight $\gamma$, except for plasma, that decreases in metrics with increasing $\gamma$ (Figure S3). Sil-score and CHI have opposite trends, deteriorating with larger $L$ and $\gamma$. To balance clustering and classification, we set $L = 3$ and $\gamma = 1$. The number of clusters in the consensus approach was fixed at six based on the consensus matrix graph (Figure S2).

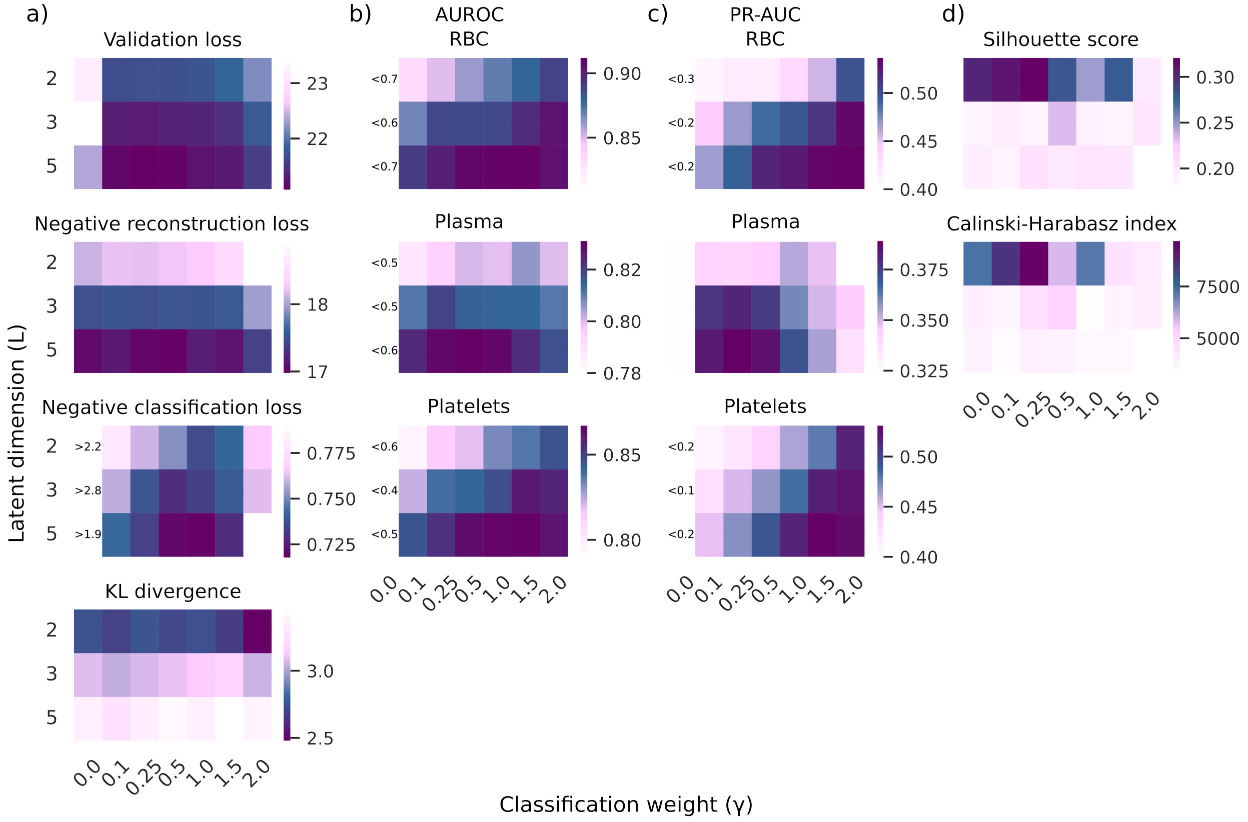

**Figure S3**: Results for grid search. a) Loss terms where clustering loss includes terms 3-6, b) AUROC of classification labels, c) PR-AUC of classification labels and d) clustering metrics.

#### S2.2 Cluster characteristics

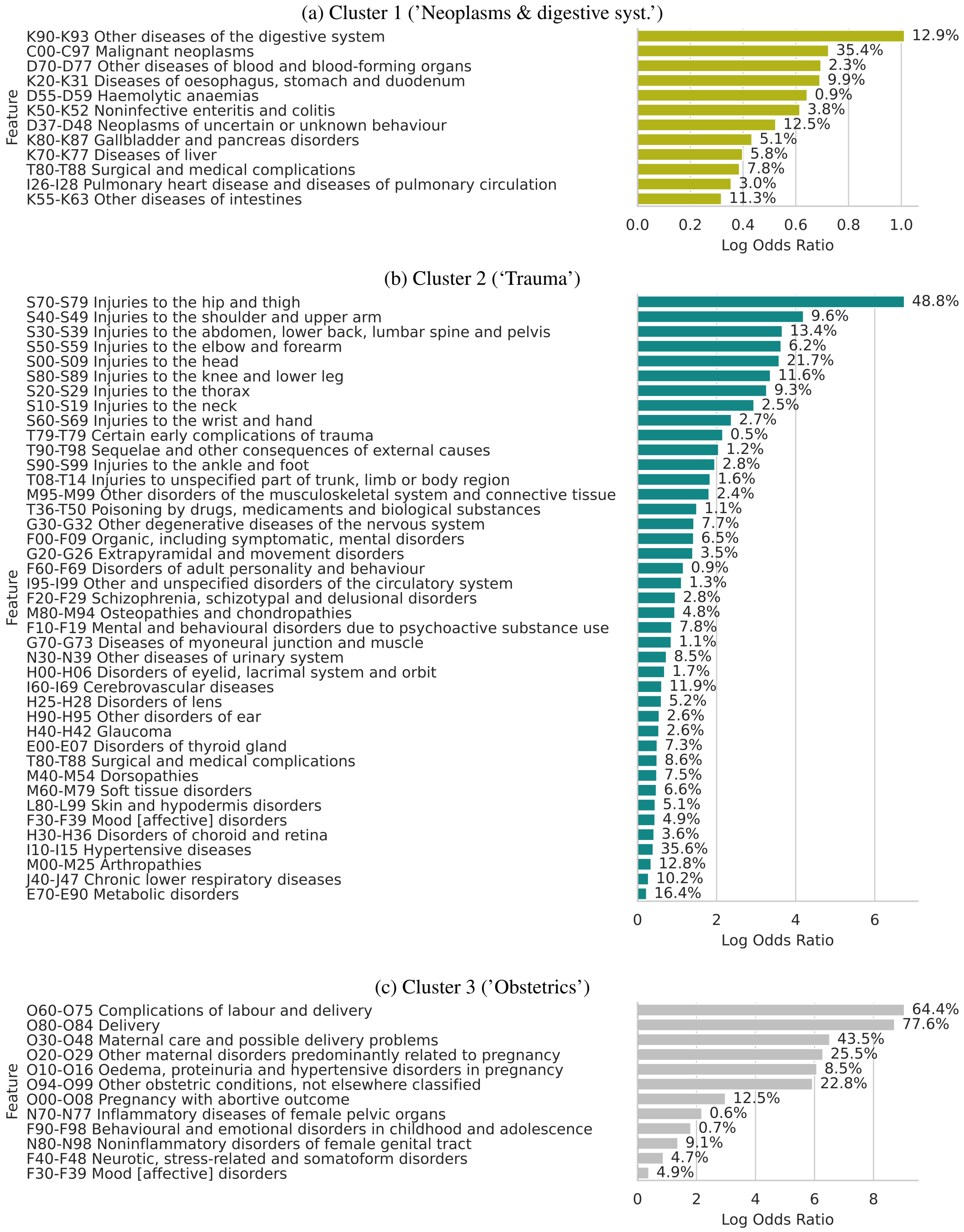

**Figure S4**: ICD level 2 categories enriched in clusters.

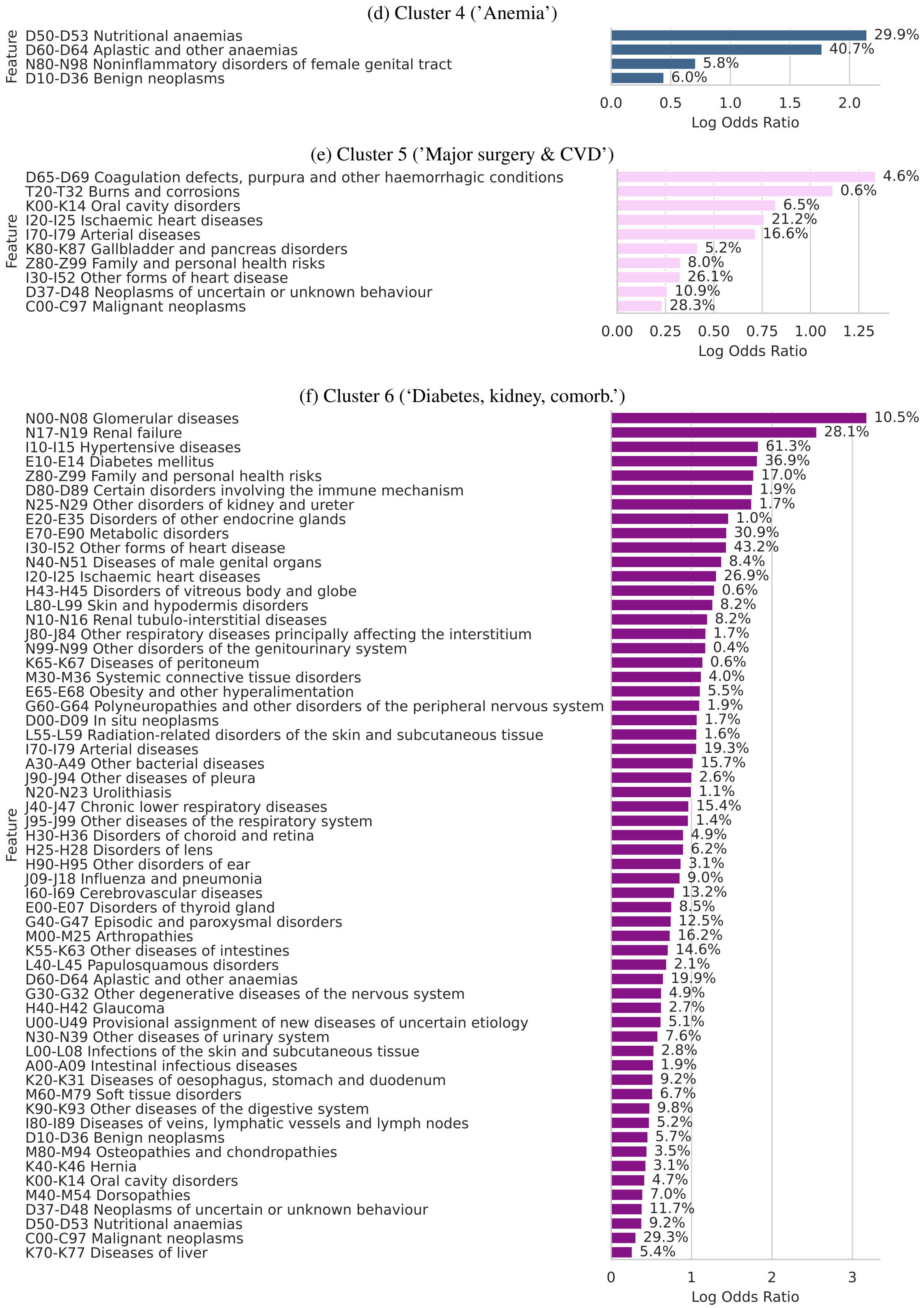

**Figure S4**: ICD level 2 categories enriched in clusters. (cont.)

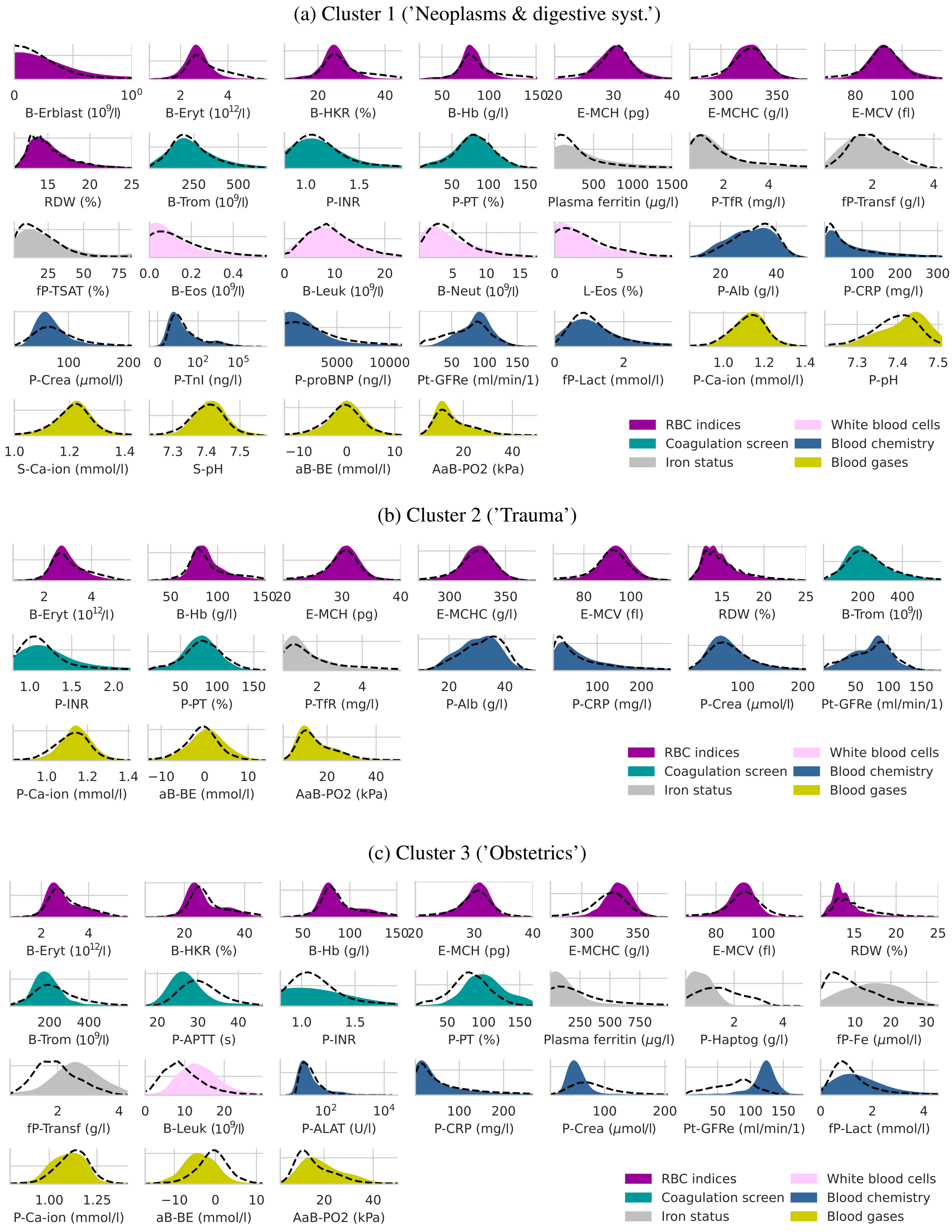

**Figure S5**: Statistically significant laboratory results of clusters.

(d) Cluster 4 ('Anemia')

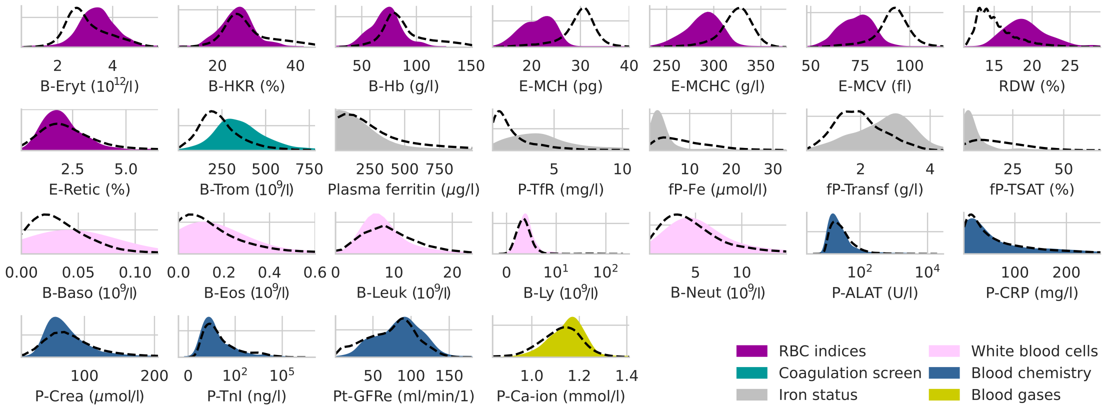

(e) Cluster 5 ('Major surgery & CVD')

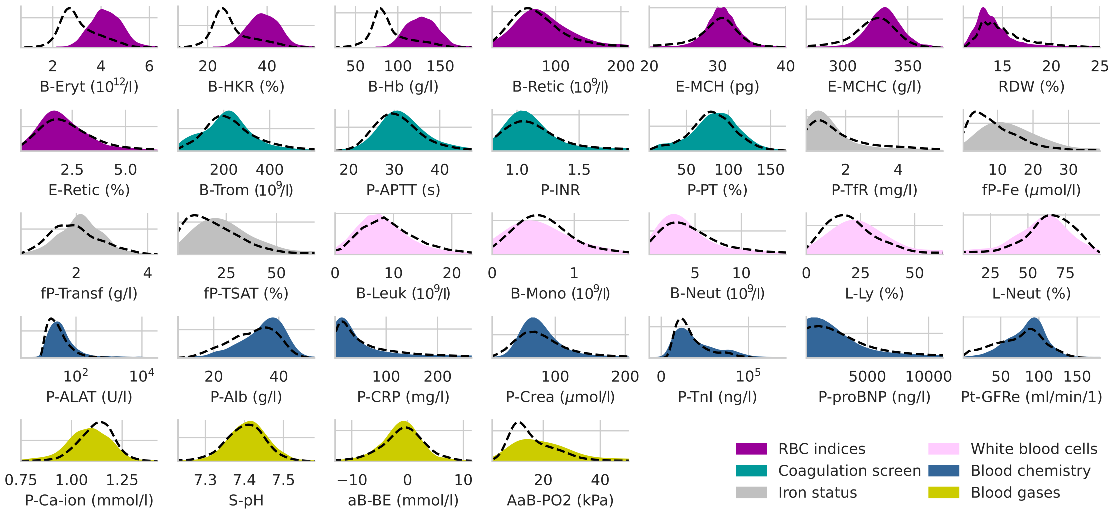

(f) Cluster 6 ('Diabetes, kidney, comorb.')

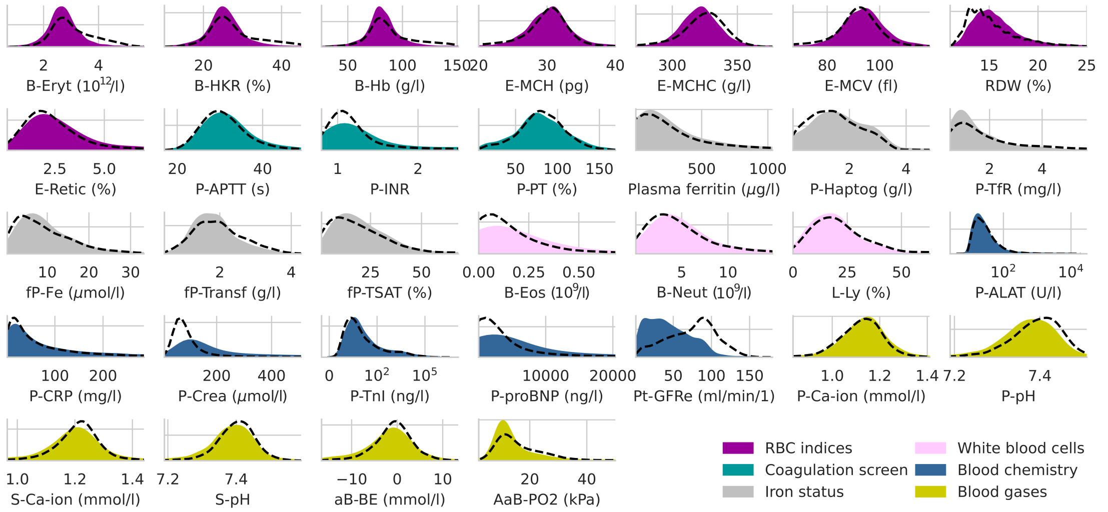

**Figure S5**: Statistically significant laboratory results of clusters. (cont.)

**Table S3**: Cluster characteristics.

| Feature | Statistic | Cluster | | | | | |
|---|---|---|---|---|---|---|---|
| | | 1 | 2 | 3 | 4 | 5 | 6 |
| Age (years) | Patients with data | 3844 | 3111 | 1597 | 1426 | 2441 | 3285 |
| | Q1 | 55 | 67 | 28 | 48 | 55 | 67 |
| | Median | 67 | 78 | 32 | 68 | 65 | 75 |
| | Q3 | 76 | 86 | 36 | 78 | 73 | 83 |
| Pre-transf bleeding (ml) | Patients with data | 1977 | 2324 | 1495 | 506 | 1948 | 1563 |
| | Q1 | 0 | 0 | 0 | 0 | 0 | 0 |
| | Median | 0 | 0 | 900 | 0 | 0 | 0 |
| | Q3 | 0 | 50 | 1650 | 0 | 600 | 0 |
| RBC units | Recipient count | 3743 | 2953 | 1541 | 1423 | 1519 | 3211 |
| | Q1 (recipients) | 1 | 1 | 1 | 1 | 1 | 1 |
| | Median (recipients) | 2 | 2 | 2 | 2 | 2 | 1 |
| | Q3 (recipients) | 2 | 2 | 2 | 2 | 2 | 2 |
| Plasma units | Recipient count | 188 | 192 | 248 | 20 | 822 | 122 |
| | Q1 (recipients) | 2 | 2 | 2 | - | 2 | 2 |
| | Median (recipients) | 2 | 2 | 2 | - | 3 | 3 |
| | Q3 (recipients) | 4 | 3 | 4 | - | 4 | 6.75 |
| Platelet units | Recipient count | 183 | 192 | 92 | 5 | 1039 | 111 |
| | Q1 (recipients) | 1 | 1 | 1 | - | 2 | 1 |
| | Median (recipients) | 2 | 2 | 2 | - | 2 | 2 |
| | Q3 (recipients) | 2 | 2 | 2 | - | 2 | 2 |
| Multicomponent transfusions (N) | RBC | >3510 | 2755 | >1290 | 1403 | 919 | >3075 |
| | Plasma | 26 | 41 | 17 | <5 | 232 | 29 |
| | Platelets | 74 | 111 | 38 | <5 | 598 | 44 |
| | RBC + Plasma | 121 | 123 | 193 | 17 | 251 | 65 |
| | RBC + Platelets | 68 | 53 | 16 | <5 | 102 | 39 |
| | Plasma + Platelets | <5 | 6 | <5 | <5 | 92 | <5 |
| | RBC+Plasma+Platelets | 40 | 22 | 37 | <5 | 247 | 27 |
| Multicomponent transfusions (%) | RBC | >91.31 | 88.56 | >80.77 | 98.39 | 37.65 | >93.6 |
| | Plasma | 0.68 | 1.32 | 1.06 | <0.36 | 9.5 | 0.88 |
| | Platelets | 1.93 | 3.57 | 2.38 | <0.36 | 24.50 | 1.34 |
| | RBC + Plasma | 3.15 | 3.95 | 12.09 | 1.19 | 10.28 | 1.98 |
| | RBC + Platelets | 1.77 | 1.70 | 1.00 | <0.36 | 4.18 | 1.19 |
| | Plasma + Platelets | <0.14 | 0.19 | <0.32 | <0.36 | 3.77 | <0.16 |
| | RBC+Plasma+Platelets | 1.04 | 0.71 | 2.32 | <0.36 | 10.12 | 0.82 |

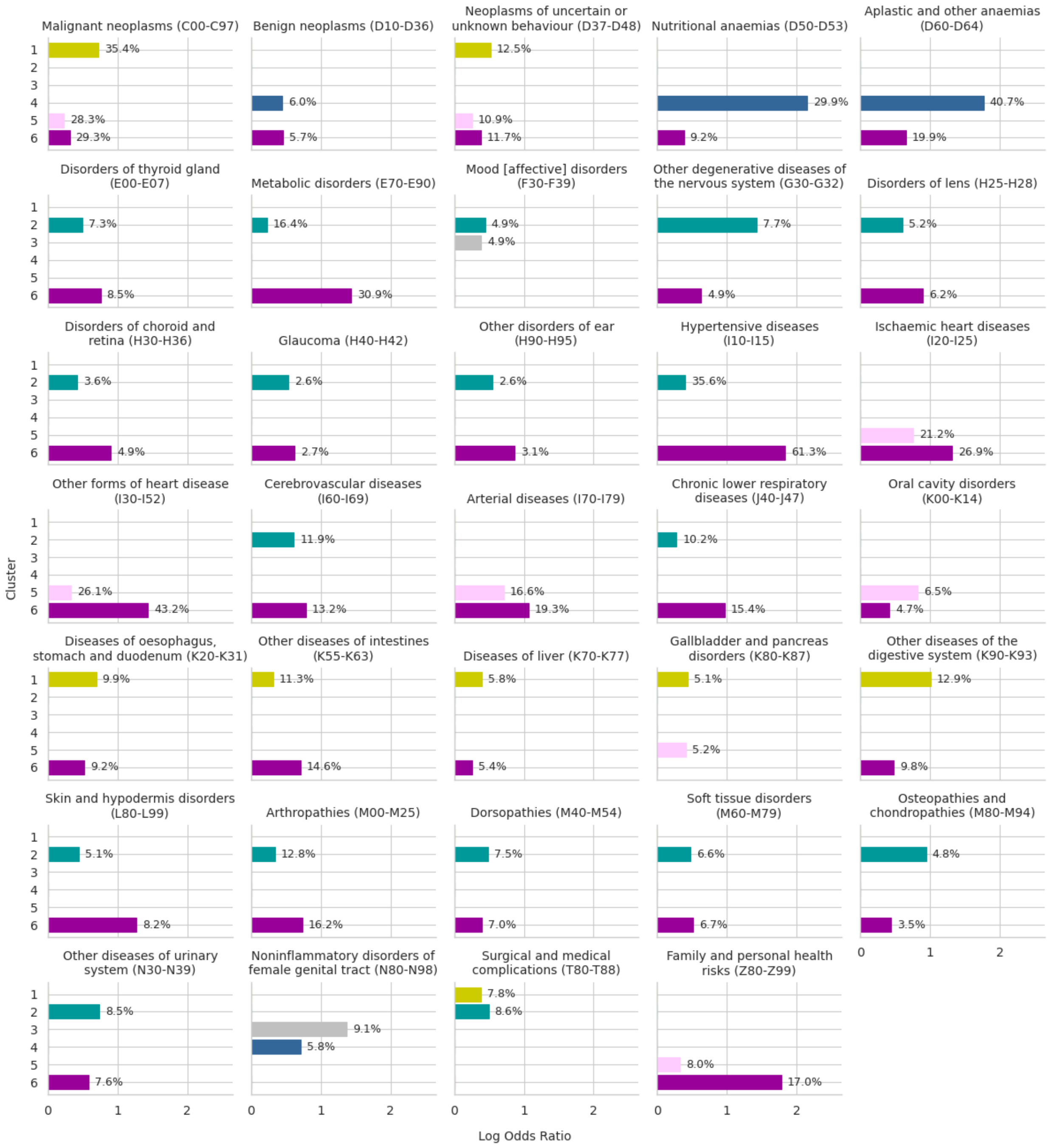

**Figure S6**: ICD level 2 categories enriched in more than one cluster.

##### S2.3 Consensus clustering results

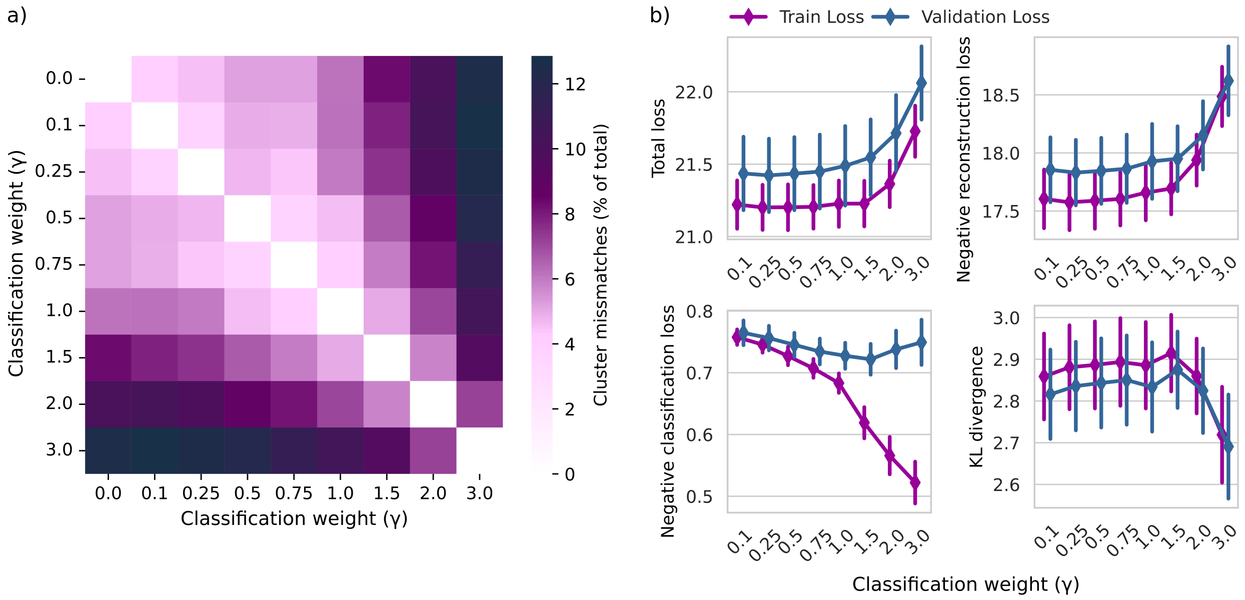

**Figure S7**: Comparison of consensus runs over different $\gamma$ a) Percent of mismatched patients between pairwise comparisons and b) mean $\pm 95\%$ confidence-interval of total loss (negative augmented ELBO) and three loss terms.

**Table S4**: Mean and standard deviation of loss terms over 80 consensus runs with varying classification weight $\gamma$.

| $\gamma$ | Early stopping at epoch | Best epoch | Total loss | | Negative reconstruction loss | | KL divergence | | Negative classification loss | |
|---|---|---|---|---|---|---|---|---|---|---|
| | | | train | val | train | val | train | val | train | val |
| 0 | $229 \pm 42$ | $180 \pm 42$ | $22.9 \pm 0.43$ | $23.1 \pm 0.45$ | $17.59 \pm 0.24$ | $17.84 \pm 0.3$ | $2.87 \pm 0.1$ | $2.83 \pm 0.11$ | $2.44 \pm 0.43$ | $2.43 \pm 0.42$ |
| 0.1 | $230 \pm 39$ | $181 \pm 39$ | $21.22 \pm 0.17$ | $21.44 \pm 0.25$ | $17.6 \pm 0.25$ | $17.86 \pm 0.28$ | $2.86 \pm 0.1$ | $2.82 \pm 0.11$ | $0.76 \pm 0.01$ | $0.76 \pm 0.02$ |
| 0.25 | $237 \pm 46$ | $188 \pm 46$ | $21.2 \pm 0.16$ | $21.42 \pm 0.25$ | $17.58 \pm 0.24$ | $17.83 \pm 0.28$ | $2.88 \pm 0.1$ | $2.84 \pm 0.11$ | $0.75 \pm 0.01$ | $0.76 \pm 0.02$ |
| 0.5 | $232 \pm 38$ | $184 \pm 38$ | $21.2 \pm 0.16$ | $21.43 \pm 0.25$ | $17.59 \pm 0.24$ | $17.85 \pm 0.28$ | $2.89 \pm 0.1$ | $2.84 \pm 0.11$ | $0.73 \pm 0.01$ | $0.75 \pm 0.02$ |
| 0.75 | $233 \pm 43$ | $183 \pm 43$ | $21.21 \pm 0.15$ | $21.45 \pm 0.26$ | $17.6 \pm 0.23$ | $17.86 \pm 0.29$ | $2.89 \pm 0.11$ | $2.85 \pm 0.11$ | $0.71 \pm 0.02$ | $0.73 \pm 0.02$ |
| 1 | $231 \pm 40$ | $182 \pm 40$ | $21.23 \pm 0.16$ | $21.49 \pm 0.27$ | $17.66 \pm 0.24$ | $17.93 \pm 0.32$ | $2.89 \pm 0.1$ | $2.83 \pm 0.11$ | $0.68 \pm 0.02$ | $0.73 \pm 0.02$ |
| 1.5 | $222 \pm 35$ | $173 \pm 34$ | $21.23 \pm 0.16$ | $21.55 \pm 0.26$ | $17.69 \pm 0.22$ | $17.95 \pm 0.28$ | $2.91 \pm 0.09$ | $2.87 \pm 0.09$ | $0.62 \pm 0.03$ | $0.72 \pm 0.02$ |
| 2 | $181 \pm 32$ | $131 \pm 32$ | $21.36 \pm 0.16$ | $21.71 \pm 0.26$ | $17.94 \pm 0.22$ | $18.15 \pm 0.29$ | $2.86 \pm 0.09$ | $2.82 \pm 0.1$ | $0.57 \pm 0.03$ | $0.74 \pm 0.03$ |
| 3 | $117 \pm 19$ | $68 \pm 18$ | $21.73 \pm 0.18$ | $22.06 \pm 0.25$ | $18.49 \pm 0.26$ | $18.62 \pm 0.3$ | $2.72 \pm 0.11$ | $2.69 \pm 0.12$ | $0.52 \pm 0.03$ | $0.75 \pm 0.04$ |

**Table S5**: Mean and standard deviation of classification metrics of plasma classification over 80 consensus runs with varying classification weight $\gamma$.

| $\gamma$ | AUROC (Plasma) | | PR-AUC (Plasma) | | PPV (Plasma) | |
|---|---|---|---|---|---|---|
| | train + val | test | train + val | test | train + val | test |
| 0 | $0.489 \pm 0.15$ | $0.489 \pm 0.139$ | $0.117 \pm 0.051$ | $0.118 \pm 0.05$ | $0.476 \pm 0.321$ | $0.475 \pm 0.316$ |
| 0.1 | $0.804 \pm 0.02$ | $0.801 \pm 0.009$ | $0.351 \pm 0.033$ | $0.357 \pm 0.012$ | $0.013 \pm 0.016$ | $0.013 \pm 0.013$ |
| 0.25 | $0.807 \pm 0.02$ | $0.803 \pm 0.008$ | $0.355 \pm 0.032$ | $0.362 \pm 0.011$ | $0.015 \pm 0.016$ | $0.015 \pm 0.016$ |
| 0.5 | $0.811 \pm 0.018$ | $0.805 \pm 0.008$ | $0.36 \pm 0.032$ | $0.361 \pm 0.01$ | $0.022 \pm 0.021$ | $0.019 \pm 0.018$ |
| 0.75 | $0.814 \pm 0.019$ | $0.806 \pm 0.008$ | $0.364 \pm 0.035$ | $0.36 \pm 0.01$ | $0.031 \pm 0.026$ | $0.026 \pm 0.02$ |
| 1 | $0.816 \pm 0.018$ | $0.806 \pm 0.008$ | $0.364 \pm 0.033$ | $0.357 \pm 0.011$ | $0.036 \pm 0.032$ | $0.034 \pm 0.027$ |
| 1.5 | $0.816 \pm 0.021$ | $0.803 \pm 0.007$ | $0.362 \pm 0.033$ | $0.348 \pm 0.013$ | $0.047 \pm 0.036$ | $0.038 \pm 0.029$ |
| 2 | $0.812 \pm 0.02$ | $0.799 \pm 0.007$ | $0.352 \pm 0.032$ | $0.339 \pm 0.014$ | $0.046 \pm 0.036$ | $0.04 \pm 0.029$ |
| 3 | $0.811 \pm 0.019$ | $0.798 \pm 0.007$ | $0.338 \pm 0.03$ | $0.328 \pm 0.016$ | $0.036 \pm 0.034$ | $0.031 \pm 0.027$ |

**Table S6**: Mean and standard deviation of classification metrics of RBC classification over 80 consensus runs with varying classification weight $\gamma$.

| $\gamma$ | AUROC (RBC) | | PR-AUC (RBC) | | PPV (RBC) | |
|---|---|---|---|---|---|---|
| | train + val | test | train + val | test | train + val | test |
| 0 | $0.525 \pm 0.183$ | $0.522 \pm 0.165$ | $0.122 \pm 0.09$ | $0.112 \pm 0.071$ | $0.481 \pm 0.275$ | $0.482 \pm 0.275$ |
| 0.1 | $0.863 \pm 0.019$ | $0.853 \pm 0.011$ | $0.43 \pm 0.042$ | $0.38 \pm 0.016$ | $0.997 \pm 0.003$ | $0.996 \pm 0.003$ |
| 0.25 | $0.87 \pm 0.019$ | $0.86 \pm 0.011$ | $0.443 \pm 0.042$ | $0.394 \pm 0.017$ | $0.995 \pm 0.004$ | $0.995 \pm 0.004$ |
| 0.5 | $0.876 \pm 0.018$ | $0.869 \pm 0.009$ | $0.452 \pm 0.046$ | $0.413 \pm 0.018$ | $0.994 \pm 0.004$ | $0.993 \pm 0.004$ |
| 0.75 | $0.882 \pm 0.018$ | $0.875 \pm 0.01$ | $0.465 \pm 0.046$ | $0.433 \pm 0.022$ | $0.991 \pm 0.004$ | $0.991 \pm 0.004$ |
| 1 | $0.885 \pm 0.017$ | $0.879 \pm 0.009$ | $0.472 \pm 0.048$ | $0.448 \pm 0.024$ | $0.987 \pm 0.006$ | $0.988 \pm 0.005$ |
| 1.5 | $0.892 \pm 0.017$ | $0.886 \pm 0.009$ | $0.489 \pm 0.048$ | $0.472 \pm 0.022$ | $0.978 \pm 0.006$ | $0.978 \pm 0.005$ |
| 2 | $0.892 \pm 0.016$ | $0.887 \pm 0.01$ | $0.495 \pm 0.046$ | $0.482 \pm 0.019$ | $0.97 \pm 0.006$ | $0.971 \pm 0.005$ |
| 3 | $0.898 \pm 0.015$ | $0.891 \pm 0.008$ | $0.509 \pm 0.049$ | $0.489 \pm 0.022$ | $0.966 \pm 0.006$ | $0.966 \pm 0.005$ |

**Table S7**: Mean and standard deviation of classification metrics of platelet classification over 80 consensus runs with varying classification weight $\gamma$.

| $\gamma$ | AUROC (Platelets) | | PR-AUC (Platelets) | | PPV (Platelets) | |
|---|---|---|---|---|---|---|
| | train + val | test | train + val | test | train + val | test |
| 0 | $0.503 \pm 0.159$ | $0.502 \pm 0.152$ | $0.132 \pm 0.086$ | $0.127 \pm 0.075$ | $0.512 \pm 0.343$ | $0.517 \pm 0.341$ |
| 0.1 | $0.828 \pm 0.02$ | $0.837 \pm 0.009$ | $0.431 \pm 0.04$ | $0.424 \pm 0.019$ | $0.125 \pm 0.059$ | $0.123 \pm 0.058$ |
| 0.25 | $0.834 \pm 0.02$ | $0.843 \pm 0.011$ | $0.442 \pm 0.04$ | $0.438 \pm 0.02$ | $0.151 \pm 0.061$ | $0.147 \pm 0.059$ |
| 0.5 | $0.841 \pm 0.019$ | $0.851 \pm 0.008$ | $0.453 \pm 0.041$ | $0.458 \pm 0.02$ | $0.185 \pm 0.063$ | $0.185 \pm 0.066$ |
| 0.75 | $0.846 \pm 0.019$ | $0.857 \pm 0.009$ | $0.465 \pm 0.039$ | $0.478 \pm 0.021$ | $0.228 \pm 0.052$ | $0.232 \pm 0.053$ |
| 1 | $0.851 \pm 0.016$ | $0.861 \pm 0.007$ | $0.476 \pm 0.039$ | $0.492 \pm 0.021$ | $0.278 \pm 0.054$ | $0.28 \pm 0.049$ |
| 1.5 | $0.859 \pm 0.017$ | $0.867 \pm 0.007$ | $0.491 \pm 0.041$ | $0.515 \pm 0.019$ | $0.369 \pm 0.049$ | $0.375 \pm 0.04$ |
| 2 | $0.857 \pm 0.018$ | $0.866 \pm 0.009$ | $0.5 \pm 0.042$ | $0.521 \pm 0.019$ | $0.415 \pm 0.045$ | $0.422 \pm 0.029$ |
| 3 | $0.863 \pm 0.017$ | $0.868 \pm 0.008$ | $0.513 \pm 0.043$ | $0.528 \pm 0.021$ | $0.446 \pm 0.042$ | $0.453 \pm 0.027$ |

**Table S8**: Mean and standard deviation of clustering metrics over 80 consensus runs with varying classification weight $\gamma$.

| $\gamma$ | Silhouette Score | | Calinski-Harabasz Index | |
|---|---|---|---|---|
| | train + val | test | train + val | test |
| 0 | $0.177 \pm 0.057$ | $0.177 \pm 0.059$ | $2021 \pm 557$ | $1066 \pm 287$ |
| 0.1 | $0.178 \pm 0.055$ | $0.18 \pm 0.054$ | $1976 \pm 622$ | $1041 \pm 315$ |
| 0.25 | $0.178 \pm 0.058$ | $0.18 \pm 0.056$ | $1935 \pm 533$ | $1017 \pm 271$ |
| 0.5 | $0.187 \pm 0.063$ | $0.189 \pm 0.066$ | $1987 \pm 587$ | $1053 \pm 311$ |
| 0.75 | $0.19 \pm 0.046$ | $0.192 \pm 0.048$ | $2029 \pm 616$ | $1052 \pm 321$ |
| 1 | $0.189 \pm 0.052$ | $0.189 \pm 0.055$ | $1998 \pm 581$ | $1037 \pm 293$ |
| 1.5 | $0.181 \pm 0.052$ | $0.177 \pm 0.057$ | $1890 \pm 448$ | $971 \pm 231$ |
| 2 | $0.176 \pm 0.059$ | $0.172 \pm 0.065$ | $1791 \pm 503$ | $933 \pm 280$ |
| 3 | $0.133 \pm 0.066$ | $0.13 \pm 0.066$ | $1520 \pm 520$ | $779 \pm 261$ |

#### S3 Experimental research

During development, the classifier module was initially incorporated into a Vanilla-VAE with isotropic standard Gaussian prior. It was noticed that increasing $\gamma$ had a direct effect to the formation of latent space, with hyperplanes dividing the latent space according to the classes. Thus, with increasing $\gamma$ the loss approaches the loss of a MLP classifier. However, similar behavior is not observed with GMM prior. This is due to the multimodality of the prior. Standard Gaussian prior restricts the latent space around one mode, which enables direct influence of the weight of the classification loss to guide the hyperplane formation around the mode. With multimodal prior, the latent space is drawn towards multimodal structure, and since classification labels are not equal to the clusters, the classification loss affects the formation of latent space only within the bounded GMM structure.
